# Artificial Intelligence-Driven Precision Oncology Uncovers Prognostic Significance of RTK-RAS Alterations in FOLFOX-Treated Early-Onset Colorectal Cancer

**DOI:** 10.64898/2025.12.13.25342208

**Authors:** Fernando C. Diaz, Brigette Waldrup, Francisco G. Carranza, Sophia Manjarrez, Enrique Velazquez-Villarreal

## Abstract

The incidence of early-onset colorectal cancer (EOCRC; <50 years) is rising rapidly among populations. Although alterations in the RTK-RAS signaling pathway are central to colorectal cancer (CRC) progression, their prognostic significance in FOLFOX-treated EOCRC remains poorly defined. We analyzed 2,515 CRC cases (H/L = 266; non-Hispanic White [NHW] = 2,249) stratified by ancestry, age at onset, and FOLFOX treatment status using Fisher’s exact, chi-square, and Kaplan-Meier survival analyses. We further employed the AI-HOPE and AI-HOPE-RTK-RAS conversational artificial intelligence platforms to integrate clinical, genomic, and treatment-level data through multi-parameter, natural language-driven queries. Among EO H/L patients, ERBB2 (2.7% vs. 15.4%, *p* = 0.01) and NF1 (4.1% vs. 19.2%, *p* = 0.01) mutations were significantly less frequent in FOLFOX-treated compared with untreated patients. Among LO H/L patients, NTRK2 mutations were less enriched in FOLFOX-treated cases (0.0% vs. 6.0%, *p* = 0.04). In FOLFOX-untreated EO H/L patients, MAPK3 (5.8% vs. 1.0% in EO NHW; *p* = 0.04) and NF1 (19.2% vs. 6.0% in LO NHW; *p* = 0.002) mutations were significantly enriched. Among EO NHW patients, IGF1R (2.1% vs. 5.3%, *p* = 0.04) and ERRFI1 (0.5% vs. 2.6%, *p*= 0.02) mutations were less frequent in FOLFOX-treated versus untreated cases. In LO NHW patients, multiple RTK-RAS genes (ERBB3, KIT, IGF1R, RET, ALK, FLT3, ERRFI1, ARAF, RAF1) were less enriched with FOLFOX exposure. Survival analyses revealed that RTK-RAS pathway alterations predicted worse overall survival in FOLFOX-untreated EO NHW patients (*p* = 0.029) but were associated with improved survival in FOLFOX-treated LO NHW patients (*p* = 0.048). Collectively, these findings indicate that RTK-RAS pathway alterations exhibit strong ancestry-, age-, and treatment-specific effects and may serve as precision biomarkers of differential chemotherapy response. The AI-enabled framework markedly accelerated integrative biomarker discovery, supporting its utility for advancing precision oncology in populations affected by EOCRC.

## 1. Introduction

Colorectal cancer (CRC) remains a major global cause of cancer morbidity and mortality, and contemporary management increasingly depends on molecular stratification to guide systemic therapy choices across the disease continuum (1–4). In parallel with advances in screening and treatment, a concerning epidemiologic shift has emerged: early-onset colorectal cancer (EOCRC), defined as diagnosis before age 50, is rising in multiple populations, expanding the clinical urgency to understand whether tumor biology, and treatment response, differs by age at onset and ancestry (1,3). Because EOCRC patients frequently present with distinct clinical trajectories and longer life-years at risk from treatment-related toxicities, identifying biomarkers that inform chemotherapy benefit versus harm is particularly important in this setting (1–3).

A central axis in CRC oncogenesis and therapy selection is the receptor tyrosine kinase-RAS signaling cascade (RTK-RAS), encompassing upstream receptor alterations (e.g., ERBB family, IGF1R, KIT), negative regulators (e.g., NF1, ERRFI1), and downstream RAF-MAPK effectors. Alterations in KRAS/NRAS/BRAF are routinely evaluated because RAS activation predicts lack of benefit from anti-EGFR monoclonal antibodies and strongly influences targeted-therapy sequencing in metastatic CRC (1,4,5–11). Large randomized trials established the clinical impact of RAS status when combining EGFR inhibitors with oxaliplatin-based chemotherapy (FOLFOX), demonstrating improved outcomes in RAS-wild-type disease and potential detriment in RAS-mutant subgroups (6,7,12–14). More broadly, RTK-RAS pathway dependencies and resistance circuits remain a major focus of precision medicine efforts, particularly as therapeutic options for RAS-mutant cancers expand and combination strategies evolve (3,15–19).

Despite this progress, the prognostic and predictive implications of RTK-RAS alterations in patients receiving standard cytotoxic regimens, especially FOLFOX, remain incompletely defined (20–25). Prior studies evaluating KRAS and related markers in the context of oxaliplatin/fluoropyrimidine therapy have reported heterogeneous results across settings, including adjuvant and metastatic disease, and across endpoints such as disease-free survival, progression-free survival, and overall survival (26–30). Mechanistic work further suggests that oxaliplatin/5-FU exposure may interact with KRAS-driven signaling to shape phenotypes relevant to resistance and tumor aggressiveness, underscoring the plausibility of gene-treatment interactions within the RTK-RAS axis (19,25,32). However, most available evidence has not been designed to resolve whether RTK-RAS alterations exert age-at-onset-specific or ancestry-associated effects on outcomes after FOLFOX exposure, an important gap given the rising burden of EOCRC and the need to advance equitable precision oncology (1,3,4).

To address these limitations, we performed an integrative clinical-genomic analysis of 2,515 CRC cases (Hispanic/Latino [H/L] = 266; non-Hispanic White [NHW] = 2,249) stratified by ancestry, age at onset (EOCRC vs. late-onset CRC [LOCRC]), and FOLFOX treatment status (33–35). Using Fisher’s exact and chi-square tests for alteration frequency comparisons and Kaplan-Meier survival analyses for outcome associations, we interrogated RTK-RAS pathway alterations (36,37) as candidate biomarkers of differential chemotherapy response. We additionally leveraged AI-HOPE (38) and the pathway-specialized AI-HOPE-RTK-RAS (39) conversational artificial intelligence framework to accelerate multi-parameter integration of clinical, genomic, and treatment-level variables through natural language-driven analytic queries, enabling rapid iteration across clinically relevant subgroup definitions.

Here, we show that RTK-RAS alterations exhibit pronounced age-, ancestry-, and treatment-specific patterns, including differential enrichment of ERBB2, NF1, NTRK2, IGF1R, and ERRFI1 by FOLFOX exposure status across EO and LO strata, and divergent survival associations in key subgroups (40–45). Collectively, these findings support RTK-RAS alterations as potential precision biomarkers of chemotherapy response heterogeneity and illustrate how AI-enabled analytics can help scale rigorous, subgroup-aware biomarker discovery for EOCRC populations (46–50).

## 2. Materials and Methods

### 2.1 Study design and data acquisition

We performed a retrospective, multi-cohort precision oncology analysis integrating clinical, genomic, and treatment data from publicly available colorectal cancer (CRC) datasets accessed through the cBioPortal for Cancer Genomics. Data were obtained from three independent CRC cohorts selected for their comprehensive somatic mutation profiling and detailed therapeutic annotations: The Cancer Genome Atlas Colorectal Adenocarcinoma (TCGA, PanCancer Atlas), MSK-CHORD, and the AACR Project GENIE Biopharma Collaborative (BPC) CRC cohort.

Eligible cases included patients with histologically confirmed colon, rectal, or colorectal adenocarcinoma and available tumor sequencing data from primary or metastatic specimens. To prevent sample-level duplication, a single representative tumor sample per patient was retained based on completeness of clinical and treatment annotation. All datasets were fully de-identified and publicly accessible.

### 2.2 Ancestry assignment and age-at-onset stratification

Patient ancestry was determined using self-reported ethnicity annotations when available. Individuals were classified as H/L if designated as “Hispanic or Latino,” “Latino,” “Hispanic, NOS,” or equivalent categories. For cases lacking explicit ethnicity data, validated surname-based algorithms were applied to infer Hispanic origin. Patients not meeting H/L criteria were categorized as NHW and served as the comparator population.

Age at diagnosis was extracted from clinical metadata. EOCRC was defined as diagnosis prior to age 50, while LOCRC was defined as diagnosis at age 50 or older. All downstream analyses were explicitly stratified by ancestry (H/L vs. NHW) and age-at-onset group (EOCRC vs. LOCRC).

### 2.3 Chemotherapy exposure classification

Treatment histories were systematically reviewed to identify exposure to FOLFOX chemotherapy, defined as administration of fluorouracil (5-FU), leucovorin, and oxaliplatin either concurrently or as part of a documented treatment sequence consistent with standard clinical practice. Patients were classified as FOLFOX-treated if all three agents were recorded with overlapping or sequential treatment dates indicative of a complete regimen.

Patients lacking documentation of one or more FOLFOX components were classified as non-FOLFOX-treated. Treatment classification was performed prior to molecular stratification to avoid bias in subgroup assignment.

### 2.4 RTK-RAS pathway gene curation and alteration definition

The RTK-RAS signaling pathway was defined using a curated gene list derived from established oncogenic signaling frameworks and CRC-focused literature. The pathway included receptor tyrosine kinases (e.g., ERBB2, ERBB3, IGF1R, KIT, RET, ALK, NTRK2, FLT3), negative regulators (e.g., NF1, ERRFI1), and downstream effectors within the RAF-MAPK cascade (e.g., ARAF, RAF1, MAPK3) (Table S1).

Somatic alteration data were extracted from cBioPortal mutation calls. Only protein-altering variants, including missense, nonsense, frameshift insertions/deletions, splice-site, and start codon mutations, were retained. RTK-RAS pathway alteration status was defined dichotomously, with a patient classified as “altered” if at least one qualifying mutation was present in any pathway gene.

### 2.5 Statistical analyses

Comparisons of mutation frequencies across ancestry, age-at-onset, and treatment-defined subgroups were conducted using Fisher’s exact test or chi-square test, as appropriate based on cell counts. Continuous clinical variables were compared using nonparametric tests.

Overall survival (OS) was defined as the interval from diagnosis to death or last follow-up. Survival distributions were estimated using the Kaplan-Meier method and compared using the log-rank test. Statistical significance was defined as a two-sided p-value < 0.05. All statistical analyses were performed using R software (version 4.3.2).

### 2.6 AI-enabled integrative analysis using AI-HOPE and AI-HOPE-RTK-RAS

To facilitate high-dimensional data integration and systematically interrogate ancestry-, age-, and treatment-specific molecular patterns, we employed the AI-HOPE (38) conversational artificial intelligence framework and its pathway-specialized module, AI-HOPE-RTK-RAS (39). These AI agents are designed to integrate structured clinical variables, genomic alteration data, and treatment annotations to enable complex, multi-parameter analyses through natural language-driven queries.

The AI platforms were first used to perform an exploratory post-hoc scan of the combined CRC cohorts, prioritizing clinically actionable questions such as: (i) whether RTK-RAS alterations were differentially enriched by FOLFOX exposure within EOCRC and LOCRC subgroups; (ii) whether ancestry-specific mutation patterns emerged within treatment-defined strata; and (iii) whether RTK-RAS alteration status modified overall survival in a treatment-dependent manner.

Following this AI-guided prioritization step, AI-HOPE-RTK-RAS (39) was used to automatically identify patient subsets meeting combined criteria (ancestry, age at onset, treatment status, and pathway alteration status), generate subgroup-specific mutation frequency tables, and stratify cohorts for survival analyses. The conversational interface enabled iterative refinement of analytical parameters, minimized manual data handling, and accelerated the transition from hypothesis generation to statistically validated inference.

## 3. Results

### 3.1 Cohort composition and baseline characteristics

The study population consisted of 2,515 CRC cases with baseline demographic, clinical, and molecular features summarized in Table 1. All cases included primary tumor sequencing data and were eligible for stratification by age at onset, ancestry, and FOLFOX treatment exposure.

**Table 1.** Baseline Clinical and Demographic Characteristics of the Study Cohorts.

| Category | Characteristic | H/L, n = 266 | NHW, n = 2,249 |
| --- | --- | --- | --- |
| **Demographics** | Sex - Male | 158 (59.4%) | 1,267 (56.3%) |
|  | Sex - Female | 108 (40.6%) | 982 (43.7%) |
| **Tumor Classification** | Colon adenocarcinoma | 164 (61.7%) | 1,328 (59.0%) |
|  | Rectal adenocarcinoma | 64 (24.1%) | 646 (28.7%) |
|  | Colorectal adenocarcinoma (NOS) | 38 (14.3%) | 275 (12.2%) |
| **Disease Stage at Diagnosis** | Stage I-III | 156 (58.6%) | 1,236 (55.0%) |
|  | Stage IV | 108 (40.6%) | 1,005 (44.7%) |
|  | Not available | 2 (0.8%) | 8 (0.4%) |
| **Microsatellite Status** | MSS | 200 (75.2%) | 1,940 (86.3%) |
|  | MSI-H | 21 (7.9%) | 209 (9.3%) |
|  | Indeterminate | 10 (3.8%) | 57 (2.5%) |
|  | Not available | 35 (13.2%) | 43 (1.9%) |
| **Age at Onset & FOLFOX Exposure** | EOCRC (<50), FOLFOX-treated | 73 (27.4%) | 375 (16.7%) |
|  | LOCRC (≥50), FOLFOX-treated | 91 (34.2%) | 919 (40.9%) |
|  | EOCRC (<50), not FOLFOX-treated | 52 (19.5%) | 302 (13.4%) |
|  | LOCRC (≥50), not FOLFOX-treated | 50 (18.8%) | 653 (29.0%) |
| **Sample Characteristics** | Primary tumor sequenced | 266 (100.0%) | 2,249 (100.0%) |
| **Ethnicity Annotation** | Spanish/Hispanic NOS | 230 (86.5%) | 0 (0.0%) |
|  | Mexican (incl. Chicano) | 30 (11.3%) | 0 (0.0%) |
|  | Hispanic or Latino (specified) | 2 (0.8%) | 0 (0.0%) |
|  | Other Spanish/Hispanic | 1 (0.4%) | 0 (0.0%) |
|  | Spanish surname only | 3 (1.1%) | 0 (0.0%) |
|  | Non-Spanish / Non-Hispanic | 0 (0.0%) | 2,249 (100.0%) |

Percentages are calculated within each ancestry group. H/L indicates Hispanic/ Latino, NHW indicates Non-Hispanic White (NHW), EOCRC indicates early-onset colorectal cancer (<50 years). LOCRC indicates late-onset colorectal cancer (≥50 years). MSS, microsatellite stable; MSI-H, microsatellite instability-high.

Sex distribution was comparable across ancestry groups, with a modest male predominance in both cohorts (59.4% in H/L and 56.3% in NHW). Tumor classification was similarly distributed, with colon adenocarcinoma representing the most frequent diagnosis in both populations (61.7% H/L; 59.0% NHW), followed by rectal adenocarcinoma and colorectal adenocarcinoma not otherwise specified.

At the time of diagnosis, the majority of patients in both cohorts presented with stage I-III disease, accounting for 58.6% of H/L and 55.0% of NHW cases, respectively. Stage IV disease was observed in 40.6% of H/L patients and 44.7% of NHW patients, with minimal missing staging data in either group. Microsatellite status was predominantly microsatellite stable (MSS), although MSS tumors were less common among H/L patients (75.2%) compared with NHW patients (86.3%). A higher proportion of missing MSI annotations was noted in the H/L cohort.

Clear differences emerged when patients were stratified jointly by age at diagnosis and FOLFOX exposure. EOCRC (<50 years) treated with FOLFOX comprised a larger fraction of the H/L cohort (27.4%) than the NHW cohort (16.7%). Conversely, LOCRC (≥50 years) treated with FOLFOX was more frequent among NHW patients (40.9%) compared with H/L patients (34.2%). H/L patients also demonstrated a higher proportion of EOCRC cases that did not receive FOLFOX (19.5% vs. 13.4% in NHW), whereas untreated LOCRC cases were more prevalent in the NHW group.

As expected, ethnicity annotations confirmed complete separation between cohorts. NHW patients were uniformly classified as non-Spanish/non-Hispanic, while H/L patients were primarily annotated as Spanish/Hispanic not otherwise specified (86.5%), followed by Mexican or Chicano origin (11.3%), with minimal representation from other Hispanic/Latino subcategories.

### 3.2 Age-, Ancestry-, and Treatment-Specific Genomic Landscapes

#### 3.2.1 RTK-RAS-related clinical and genomic features in H/L patients

Among H/L patients, age at diagnosis and chemotherapy exposure were associated with measurable differences in both clinical and molecular features (Table 2a). In EOCRC, individuals treated with FOLFOX were diagnosed at a slightly older age than those not receiving FOLFOX, although this difference did not reach statistical significance. In contrast, among LOCRC H/L patients, FOLFOX-treated individuals were diagnosed at a significantly younger age than their untreated counterparts.

**Table 2.**
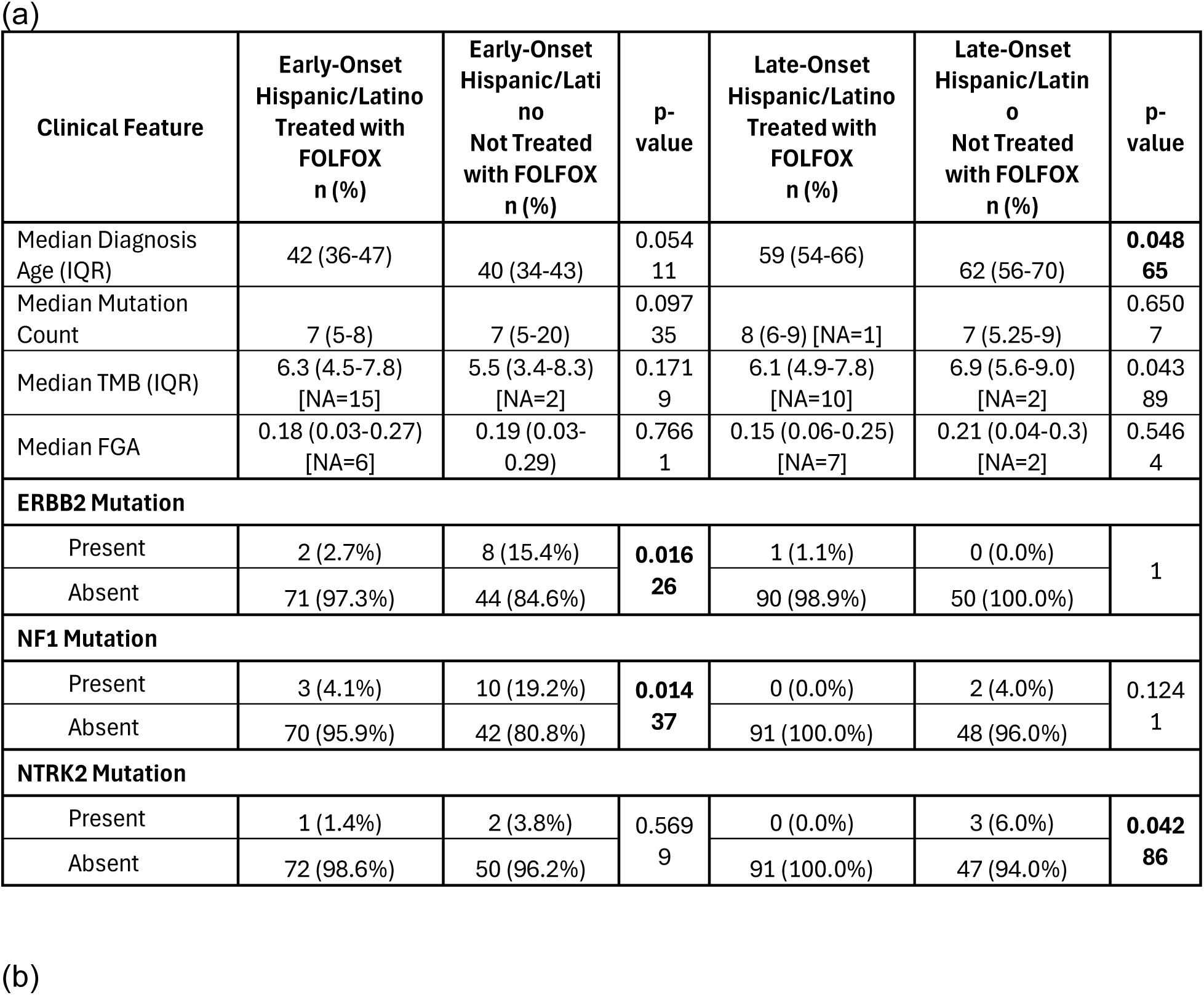
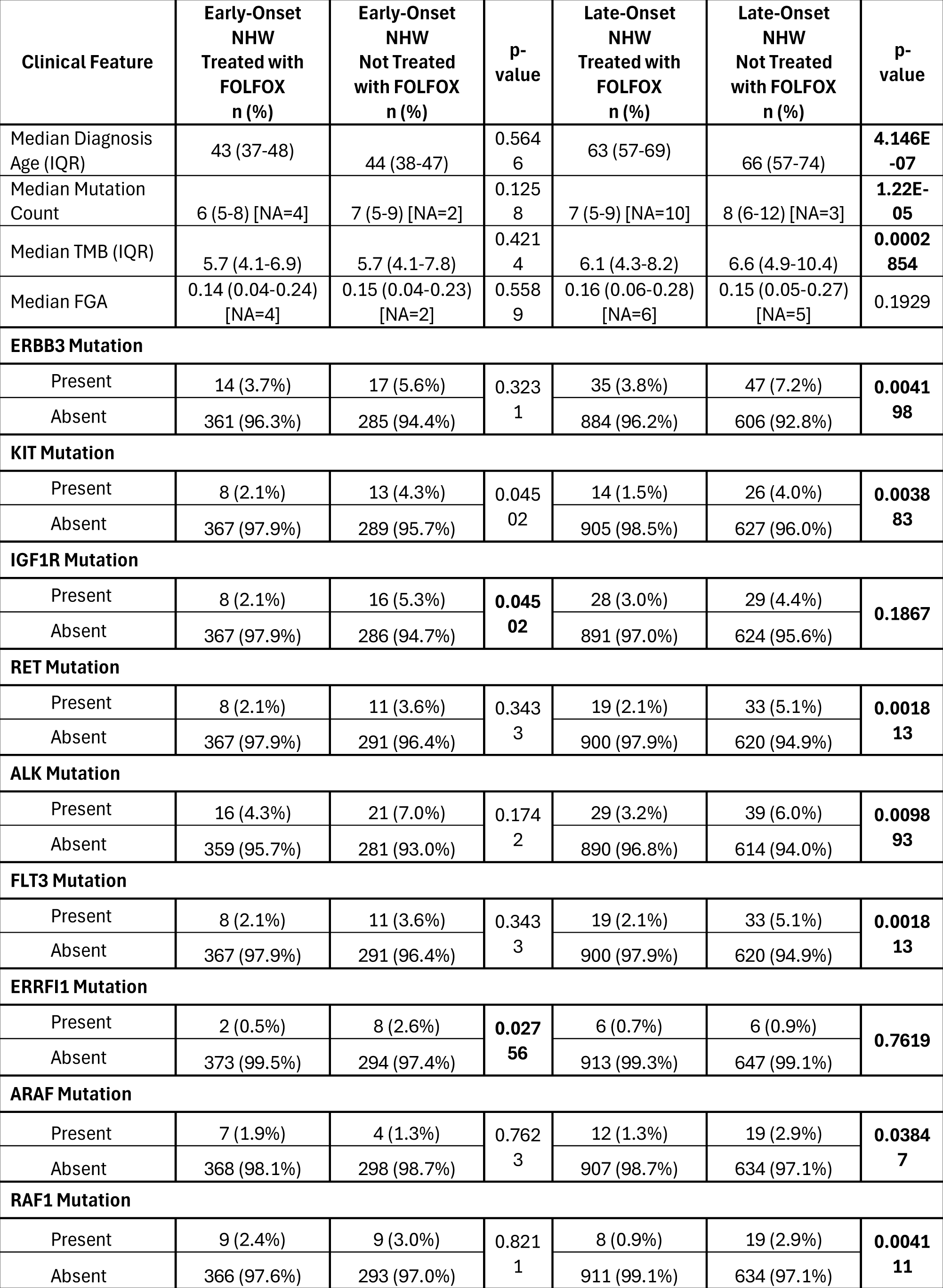
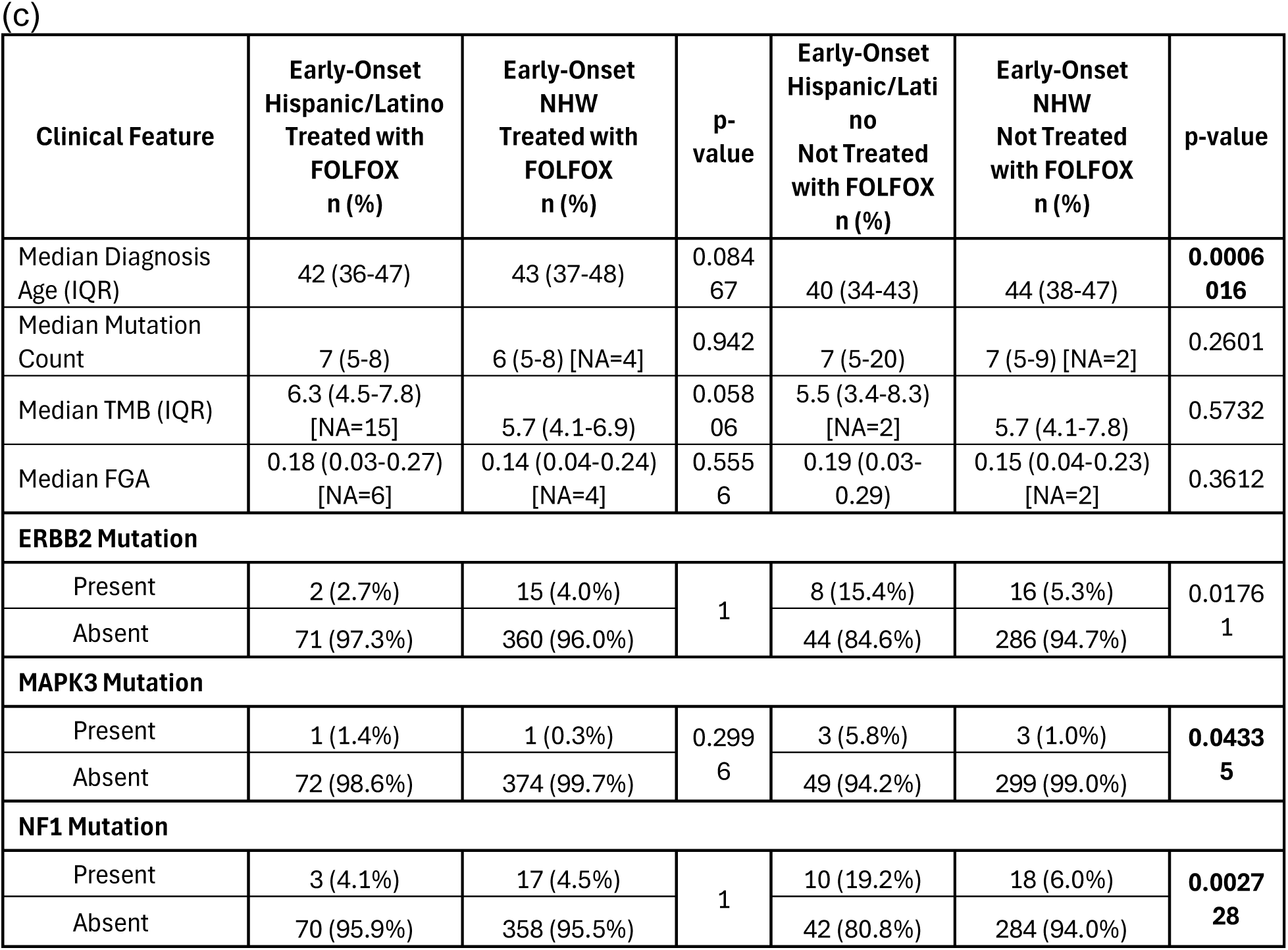
Comparative clinical, genomic, and RTK-RAS pathway features across early (EOCRC) and late (LOCRC) onset colorectal cancer subgroups stratified by ancestry and FOLFOX exposure. This table summarizes key clinical and molecular differences, including median age at diagnosis, mutation burden metrics (total mutation count, tumor mutational burden [TMB], and fraction of genome altered [FGA]), and the prevalence of selected RTK-RAS-related gene alterations, across the following comparisons: **(a)** Early-onset CRC (EOCRC) and LOCRC within Hispanic/Latino (H/L) patients, stratified by FOLFOX treatment status; **(b)** EOCRC and LOCRC within Non-Hispanic White (NHW) patients, stratified by FOLFOX treatment status; **(c)** ancestry-based comparisons between H/L and NHW patients within EOCRC, shown separately for FOLFOX-treated and non-FOLFOX-treated groups.

Global genomic metrics were largely comparable across treatment groups in EOCRC H/L patients, including mutation burden, tumor mutational burden (TMB), and fraction of genome altered (FGA). However, in LOCRC H/L disease, non-FOLFOX-treated patients exhibited significantly higher TMB compared with those receiving FOLFOX, while mutation counts and FGA remained similar.

Despite these overall similarities, treatment-associated differences emerged at the gene level. In EOCRC H/L patients, ERBB2 and NF1 mutations were significantly less frequent among those treated with FOLFOX compared with untreated patients. In LOCRC H/L patients, NTRK2 mutations were observed exclusively in the non-FOLFOX-treated group, indicating a treatment-dependent depletion of this alteration in exposed cases.

#### 3.2.2 Genomic stratification of NHW patients by age and FOLFOX exposure

Analysis of NHW patients revealed more pronounced treatment-associated genomic differences, particularly in LOCRC disease (Table 2b). Among EOCRC NHW patients, age at diagnosis and global genomic metrics, including mutation count, TMB, and FGA, did not differ significantly by FOLFOX treatment status.

In contrast, LOCRC NHW patients receiving FOLFOX were diagnosed at significantly younger ages than untreated patients and exhibited lower mutation counts and TMB. Multiple RTK-RAS pathway genes showed significantly reduced mutation frequencies in FOLFOX-treated LOCRC NHW patients, including ERBB3, KIT, RET, ALK, FLT3, ARAF, and RAF1. ERRFI1 mutations were significantly less frequent in FOLFOX-treated EOCRC NHW patients, while IGF1R mutations were less common among FOLFOX-treated EOCRC NHW patients compared with untreated cases. These findings indicate a broad, treatment-associated shift in RTK-RAS alteration profiles in NHW CRC, most evident in LOCRC disease.

#### 3.2.3 Ancestry-specific differences in EOCRC

Direct comparison of EOCRC H/L and NHW patients highlighted ancestry-associated differences that were modified by treatment exposure (Table 2c). Among FOLFOX-treated EOCRC patients, age at diagnosis, mutation burden, TMB, and FGA were similar between ancestry groups. However, among EOCRC patients not treated with FOLFOX, H/L individuals were diagnosed at significantly younger ages than NHW patients.

At the gene level, ERBB2 mutations were significantly more frequent in untreated EOCRC H/L patients compared with untreated NHW patients. Additionally, MAPK3 and NF1 mutations were enriched in untreated EOCRC H/L patients relative to their NHW counterparts, whereas no significant ancestry-related differences were observed among FOLFOX-treated EOCRC patients.

### 3.3 RTK-RAS Pathway Alterations by Age at Onset, Ancestry, and FOLFOX Treatment Status

The distribution of RTK-RAS pathway alterations was evaluated across H/L and NHW CRC cohorts, stratified by age at diagnosis and exposure to FOLFOX chemotherapy (Tables 3a-3d).

**Table 3.** Distribution of RTK-RAS pathway alteration status across colorectal cancer (CRC) subgroups defined by age at diagnosis, ancestry, and FOLFOX exposure. This table presents pathway-level frequencies of RTK-RAS alterations (present vs. absent) among CRC patients stratified by early-onset (EOCRC, <50 years) and LOCRC (≥50 years) disease, Hispanic/Latino (H/L) versus non-Hispanic White (NHW) ancestry, and receipt of FOLFOX chemotherapy. Sub-analyses include: (a) treatment-stratified comparisons within early- and LOCRC H/L patients; (b) treatment-stratified comparisons within EOCRC and LOCRC NHW patients; (c) ancestry-based comparisons among EOCRC patients by FOLFOX treatment status; and (d) ancestry-based comparisons among LOCRC patients by FOLFOX treatment status. Reported values reflect the proportion of patients harboring at least one RTK-RAS pathway alteration within each subgroup, with statistical comparisons performed using chi-square or Fisher’s exact tests, as appropriate.

| Pathway Alterations | Early-Onset<br>Hispanic/Lati<br>no<br>Treated with<br>FOLFOX<br>n (%) | Early-Onset<br>Hispanic/Lati<br>no<br>Not Treated<br>with FOLFOX<br>n (%) | p-value | Late-Onset<br>Hispanic/Lati<br>no<br>Treated with<br>FOLFOX<br>n (%) | Late-Onset<br>Hispanic/Lati<br>no<br>Not Treated<br>with FOLFOX<br>n (%) | p-value |
| --- | --- | --- | --- | --- | --- | --- |
| RTK/RAS Alterations Present | 47 (64.4%) | 35 (67.3%) | 0.8822 | 70 (76.9%) | 38 (76.0%) | 1 |
| RTK/RAS Alterations Absent | 26 (35.6%) | 17 (32.7%) |  | 21 (23.1%) | 12 (24.0%) |  |

(b)
| Pathway Alterations | Early-Onset<br>NHW<br>Treated with<br>FOLFOX<br>n (%) | Early-Onset<br>NHW<br>Not Treated<br>with FOLFOX<br>n (%) | p-value | Late-Onset<br>NHW<br>Treated with<br>FOLFOX<br>n (%) | Late-Onset<br>NHW<br>Not Treated<br>with FOLFOX<br>n (%) | p-value |
| --- | --- | --- | --- | --- | --- | --- |
| RTK/RAS Alterations Present | 249 (66.4%) | 185 (61.3%) | 0.1916 | 639 (69.5%) | 459 (70.3%) | 0.7892 |
| RTK/RAS Alterations Absent | 126 (33.6%) | 117 (38.7%) |  | 280 (30.5%) | 194 (29.7%) |  |

(c)
| Pathway Alterations | Early-Onset<br>Hispanic/Latin<br>o<br>Treated with<br>FOLFOX<br>n (%) | Early-Onset<br>NHW<br>Treated with<br>FOLFOX<br>n (%) | p-value | Early-Onset<br>Hispanic/Latin<br>o<br>Not Treated<br>with FOLFOX<br>n (%) | Early-Onset<br>NHW<br>Not Treated<br>with FOLFOX<br>n (%) | p-value |
| --- | --- | --- | --- | --- | --- | --- |
| RTK/RAS Alterations Present | 47 (64.4%) | 249 (66.4%) | 0.8432 | 35 (67.3%) | 185 (61.3%) | 0.4991 |
| RTK/RAS Alterations Absent | 26 (35.6%) | 126 (33.6%) |  | 17 (32.7%) | 117 (38.7%) |  |

(d)
| Pathway Alterations | Late-Onset<br>Hispanic/Latin<br>o<br>Treated with<br>FOLFOX<br>n (%) | Late-Onset<br>NHW<br>Treated with<br>FOLFOX<br>n (%) | p-value | Late-Onset<br>Hispanic/Latin<br>o<br>Not Treated<br>with FOLFOX<br>n (%) | Late-Onset<br>NHW<br>Not Treated<br>with FOLFOX<br>n (%) | p-value |
| --- | --- | --- | --- | --- | --- | --- |
| RTK/RAS Alterations Present | 70 (76.9%) | 639 (69.5%) | 0.1769 | 38 (76.0%) | 459 (70.3%) | 0.4879 |
| RTK/RAS Alterations Absent | 21 (23.1%) | 280 (30.5%) |  | 12 (24.0%) | 194 (29.7%) |  |

Within the H/L cohort, RTK-RAS pathway alteration frequencies were high and did not differ significantly by treatment status in either age group. Among EOCRC H/L patients, alterations were detected in 64.4% of FOLFOX-treated cases and 67.3% of non-FOLFOX-treated cases (p = 0.8822). Similarly, in LOCRC H/L patients, alteration prevalence was nearly identical between FOLFOX-treated (76.9%) and untreated patients (76.0%; p = 1.0), indicating no measurable treatment-associated difference in pathway-level alteration frequency within this ancestry group.

In the NHW cohort, RTK-RAS alterations were also common across all strata. Among EOCRC NHW patients, pathway alterations were observed in 66.4% of FOLFOX-treated cases and 61.3% of non-treated cases (p = 0.1916). In LOCRC NHW patients, alteration prevalence was comparable between FOLFOX-treated (69.5%) and non-treated groups (70.3%; p = 0.7892), demonstrating stability of pathway-level frequencies regardless of treatment exposure.

Between-ancestry comparisons in EOCRC disease showed no significant differences in RTK-RAS alteration prevalence. Among FOLFOX-treated EOCRC patients, alterations were present in 64.4% of H/L cases and 66.4% of NHW cases (p = 0.8432). In the non-FOLFOX-treated EOCRC group, alteration frequencies were similarly comparable between H/L (67.3%) and NHW (61.3%) patients (p = 0.4991).

Ancestry-based comparisons in LOCRC disease also revealed no statistically significant differences. Among FOLFOX-treated LOCRC patients, RTK-RAS alterations were detected in 76.9% of H/L cases and 69.5% of NHW cases (p = 0.1769). In non-FOLFOX-treated LOCRC patients, alteration prevalence was 76.0% in H/L patients and 70.3% in NHW patients (p = 0.4879).

Across all age, ancestry, and treatment strata, the majority of CRC patients harbored at least one RTK-RAS pathway alteration. Although overall pathway-level frequencies were largely conserved, these findings provide an essential framework for interpreting the gene-level, ancestry-specific, and treatment-dependent differences observed in subsequent analyses, underscoring the importance of dissecting RTK-RAS signaling beyond aggregate pathway prevalence.

### 3.4 Gene-level RTK-RAS alteration patterns across ancestry, age of onset, and FOLFOX exposure

Gene-specific comparisons across RTK-RAS pathway members demonstrated that most loci exhibited broadly similar mutation frequencies across strata, with only a limited subset showing statistically significant treatment- or subgroup-associated differences (Tables S1-S12).

#### 3.4.1 EOCRC H/L: FOLFOX-treated vs. not treated

Within EO H/L patients, two receptor tyrosine kinase/RAS-regulator genes differed by treatment exposure (Table S1). ERBB2alterations were less common in FOLFOX-treated EOCRC H/L cases (2.7%) than in untreated EOCRC H/L cases (15.4%; p = 0.016). Similarly, NF1 mutations were reduced in treated EO H/L patients (4.1%) relative to untreated EOCRC H/L patients (19.2%; p = 0.014). A second RTK gene signal was also observed: FGFR2 alterations were absent in treated EOCRC H/L patients (0.0%) but present in 7.7% of untreated EOCRC H/L cases (p = 0.028). Most other RTK-RAS genes, including KRAS (41.1% vs. 34.6%), NRAS, BRAF, and downstream MAPK components, did not differ significantly by FOLFOX exposure.

#### 3.4.2 LOCRC H/L: FOLFOX-treated vs. not treated

In LOCRC H/L patients, gene-level frequencies were largely stable across treatment groups (Table S2). The most notable treatment-associated difference involved NTRK2, which was absent in FOLFOX-treated LOCRC H/L cases (0.0%) but detected in 6.0% of untreated LOCRC H/L cases (p = 0.043). Other genes, including KRAS (∼41-43%) and BRAF (16.5% vs. 16.0%), were comparable between treated and untreated groups.

#### 3.4.3 Age-of-onset contrasts within H/L

When restricting to FOLFOX-treated H/L patients (EOCRC vs. LOCRC; Table S3), BRAF mutations were significantly more frequent in LOCRC disease (16.5%) than EOCRC disease (4.1%; p = 0.012), while most other genes were similar. In the non-FOLFOX H/L stratum (EO vs. LO; Table S4), NF1 alterations were enriched in EOCRC relative to LOCRC (19.2% vs. 4.0%; p = 0.028), consistent with the EOCRC-specific NF1 signal seen in treatment comparisons.

#### 3.4.4 NHW patterns by age and treatment

Across NHW cohorts, most RTK-RAS genes showed modest or no treatment-associated shifts, but EOCRC (Table S5) LOCRC (Table S6) disease demonstrated several significant differences when comparing treated vs. untreated cases. Specifically in LOCRC, ERBB3(3.8% vs. 7.2%; p = 0.004), KIT (1.5% vs. 4.0%; p = 0.004), RET (2.1% vs. 5.1%; p = 0.002), ALK (3.2% vs. 6.0%; p = 0.010), ARAF (1.3% vs. 2.9%; p = 0.038), and RAF1 (0.9% vs. 2.9%; p = 0.004) were all less frequent among LOCRC NHW patients receiving FOLFOX than among those not treated with FOLFOX. Additional LOCRC NHW differences included higher MAPK1 (0.1% vs. 0.9%; p = 0.023) and lower SOS1 (0.9% vs. 2.3%; p = 0.035) in treated vs. untreated patients. In contrast, EOCRC NHW comparisons were more limited, with BRAF higher in LOCRC vs. EOCRC among FOLFOX-treated patients (11.1% vs. 7.2%; p = 0.043; Table S7) and similarly higher in LO vs. EO among untreated patients (14.2% vs. 7.9%; p = 0.008; Table S8).

#### 3.4.5 Ancestry contrasts within EOCRC and LOCRC strata

Across EOCRC FOLFOX-treated patients (H/L vs. NHW; Table S9), no single RTK-RAS gene differed significantly by ancestry. However, among EOCRC patients not treated with FOLFOX (Table S10), multiple alterations were more frequent in H/L than NHW, including ERBB2 (15.4% vs. 5.3%; p = 0.018), MAPK3 (5.8% vs. 1.0%; p = 0.043), CBL (9.6% vs. 1.3%; p = 0.0045), and NF1 (19.2% vs. 6.0%; p = 0.0027). In LOCRC comparisons, ancestry-associated differences were limited: in FOLFOX-treated LOCRC patients, MET was more frequent in H/L than NHW (4.4% vs. 1.3%; p = 0.048; Table S11), whereas in LOCRC untreated patients NTRK2 was higher in H/L than NHW (6.0% vs. 1.4%; p = 0.047; Table S12).

These gene-level analyses indicate that while canonical drivers such as KRAS and BRAF remain prevalent across subgroups, the most prominent subgroup-specific signals involved ERBB2 and NF1 in EOCRC H/L disease (particularly among untreated patients), and multiple RTK/RAS-associated loci in LOCRC NHW patients showing reduced mutation frequencies in the FOLFOX-treated stratum.

### 3.5 Mutational landscape of the RTK-RAS pathway

#### 3.5.1 EOCRC H/L CRC

Figure 1a summarizes the RTK-RAS pathway alteration profile in EOCRC Hispanic/Latino (H/L) CRC (n = 123), integrating per-sample TMB, mutation class, and FOLFOX exposure. Overall, 82 tumors (66.7%) harbored at least one RTK-RAS pathway alteration, indicating that pathway disruption is common in this subgroup. At the gene level, KRAS was the dominant event (39%), largely comprised of missense substitutions, consistent with canonical activating variants. A second tier of recurrently altered genes included NF1 (11%) and multiple receptor tyrosine kinases, led by ERBB2 (8%), ERBB4 (8%), and ERBB3 (6%), alongside PDGFRA (6%), CBL (6%), BRAF (6%), and ROS1 (6%). Additional alterations were observed at lower frequencies across the pathway (generally ∼2-4%), including NRAS, EGFR, FGFR-family members, KIT, ALK, RET, RAF1, MAP2K1, and MAPK3, reflecting a broad but shallow tail of RTK-RAS gene involvement.

**Figure 1.**
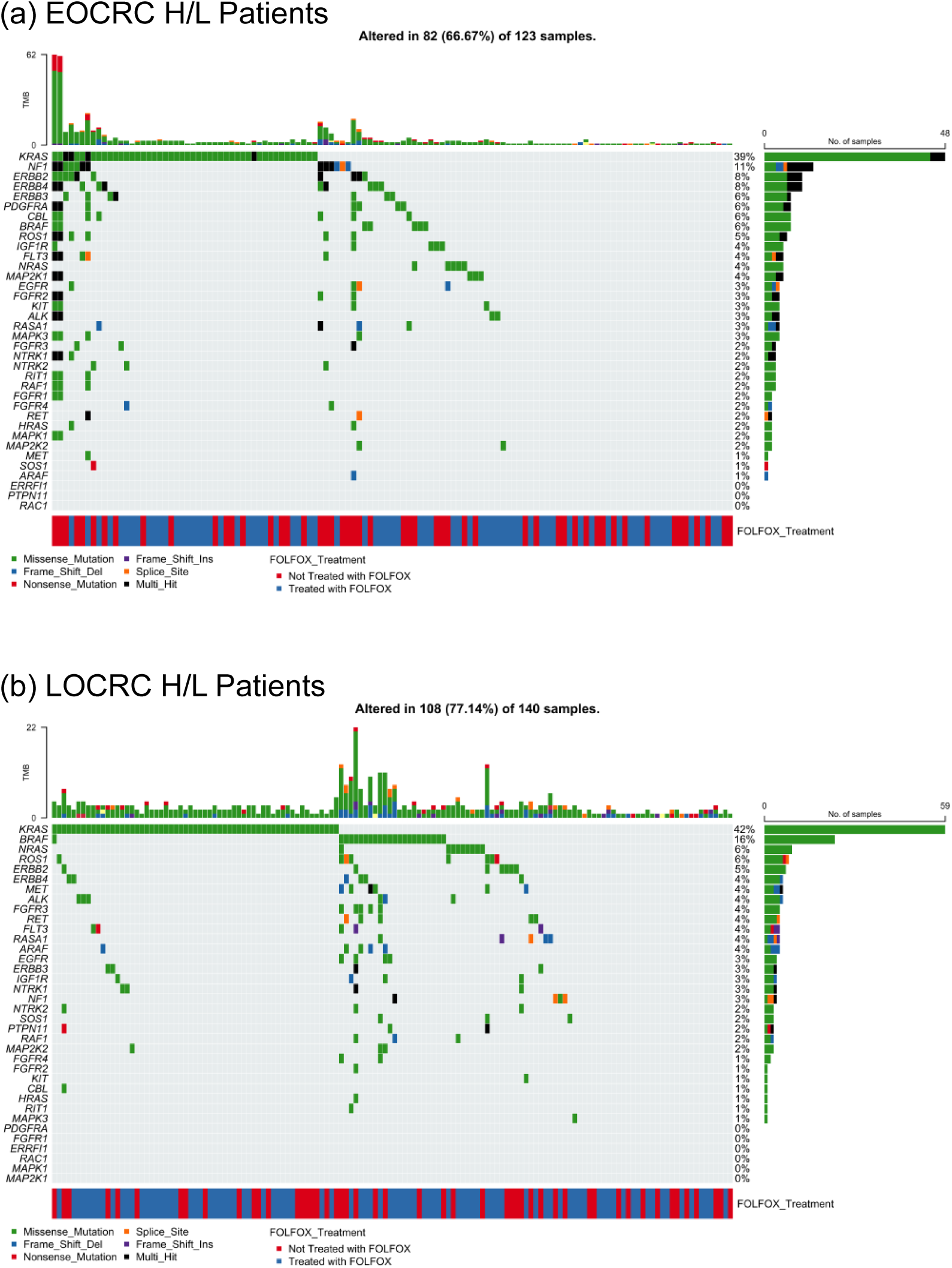
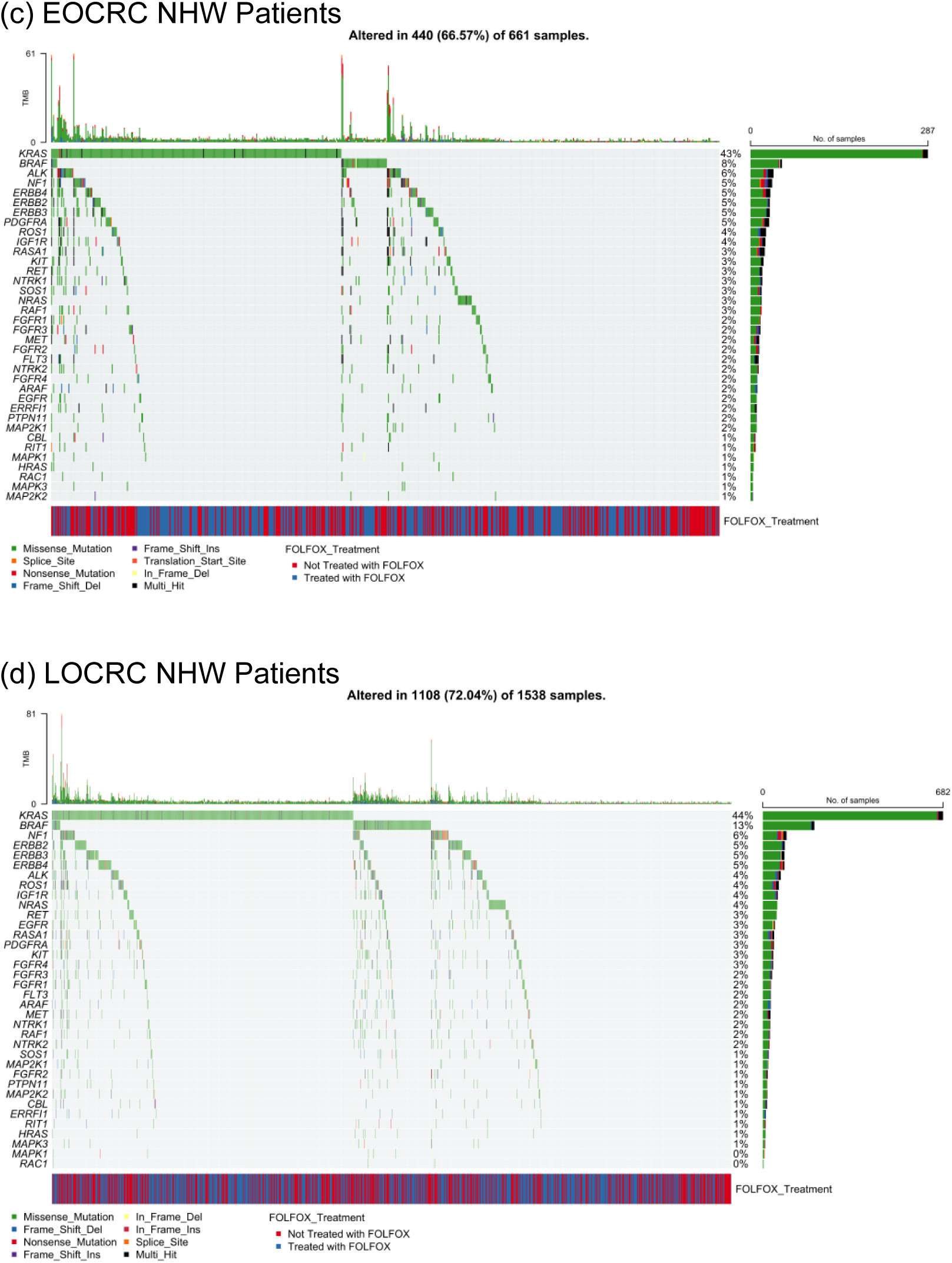
RTK-RAS pathway mutational architecture in Hispanic/Latino colorectal cancer by age at onset. Oncoprint visualizations depict somatic alterations across core RTK-RAS pathway genes in Hispanic/Latino (H/L) colorectal cancer, stratified by early-onset (EOCRC) (Figures 1a, c) and late-onset (LOCRC) (Figures 1b, d) disease. Each column represents an individual tumor, with rows corresponding to pathway genes and color-coded annotations indicating mutation classes. Upper bar plots summarize tumor mutational burden (TMB), while lower annotations denote FOLFOX treatment exposure. Side panels report gene-level alteration frequencies, enabling comparison of pathway disruption patterns and mutation spectra between EOCRC and LOCRC H/L CRC and Non-Hispanic Whites (NHW).

Across the oncoprint, alterations were predominantly missense, with intermittent truncating (frameshift/nonsense) and multi-hit patterns in select genes (Table S13). TMB values were mostly in the low-to-moderate range, with a small number of higher-TMB outliers that did not cluster within a single driver gene. Importantly, FOLFOX-treated and untreated cases were interspersed throughout the alteration spectrum, without an obvious segregation by treatment status, suggesting that the overall RTK-RAS mutational architecture in EOCRC H/L CRC is not driven by a single FOLFOX-associated genomic signature at the level of this visualization.

#### 3.5.2 LOCRC H/L CRC

The distribution of RTK–RAS pathway alterations in late-onset Hispanic/Latino colorectal cancer (LOCRC; n = 140) is shown, combining gene-level mutation frequencies with mutation classes, TMB, and FOLFOX treatment annotation. Overall, 108 tumors (77.1%) harbored at least one RTK-RAS pathway alteration, indicating a higher prevalence of pathway disruption in LOCRC compared with EOCRC H/L disease.

Consistent with canonical CRC biology, KRAS was the most frequently altered gene (42%), predominantly through missense mutations, reflecting activating hotspot variants. Additional recurrent alterations were observed in BRAF (16%) and NRAS (6%), together highlighting MAPK-axis engagement in a substantial fraction of tumors. Among receptor tyrosine kinases, ROS1 (6%), ERBB2 (5%), ERBB4 (4%), ALK (4%), FGFR3 (4%), RET (4%), and IGF1R (3%) contributed to pathway heterogeneity, while downstream regulators such as NF1 (3%), RAF1 (3%), and MAP2K1 (2%) were detected at lower frequencies.

The majority of alterations across genes consisted of missense substitutions (Table S13), with sporadic frameshift, nonsense, splice-site, and multi-hit events, particularly among RTK family members and negative regulators. TMB values were generally low to moderate across samples, with a limited number of higher-TMB outliers that did not cluster around specific RTK-RAS genes.

Assessment of treatment annotation revealed that FOLFOX-treated and untreated tumors were distributed across the full spectrum of RTK-RAS alterations, without clear segregation by gene or mutation class. This pattern suggests that, at the level of pathway-wide mutational architecture, LOCRC H/L CRC exhibits substantial RTK-RAS dysregulation that is not dominated by a single treatment-associated genomic signature, supporting subsequent analyses focused on gene-specific and survival-related effects.

#### 3.5.3 EOCRC NHW CRC

The RTK–RAS mutational landscape in early-onset non-Hispanic White (NHW) colorectal cancer (n = 661) is illustrated, displaying gene-specific alteration frequencies together with mutation types, TMB, and FOLFOX treatment status. Overall, 440 tumors (66.6%) harbored at least one RTK-RAS pathway alteration, demonstrating that RTK-RAS dysregulation is a dominant molecular feature of EOCRC in NHW patients.

As expected, KRAS was the most frequently altered gene (43%), largely through missense mutations, consistent with activating hotspot variants. Additional recurrent alterations were observed in BRAF (8%), ALK (6%), NF1 (5%), ERBB4 (5%), and ERBB2 (5%), followed by ERBB3 (5%), PDGFRA (5%), and ROS1 (4%). A long tail of less frequent events involved IGF1R, RASA1, KIT, RET, NTRK1, SOS1, NRAS, RAF1, FGFR-family members, MET, and downstream MAPK components, each occurring in approximately 1-3% of cases.

Across the cohort, alterations were predominantly missense substitutions (Table S13), with occasional frameshift, nonsense, splice-site, translation start site, and multi-hit events scattered among RTK and RAS-regulatory genes. The TMB distribution was largely low to moderate, with a subset of tumors exhibiting elevated TMB that did not map to a single dominant RTK-RAS gene, suggesting heterogeneous mutational processes rather than pathway-specific hypermutation.

Evaluation of treatment annotation showed that FOLFOX-treated and untreated EO NHW tumors were broadly intermingled across the oncoprint, without clear segregation by gene or mutation class. When considered alongside the EOCRC H/L cohort, EO NHW patients demonstrated a similarly high overall prevalence of RTK-RAS alterations but with differences in the relative contribution of specific receptor tyrosine kinases and regulatory genes, underscoring ancestry-specific variation within a shared RTK-RAS-driven oncogenic framework.

#### 3.5.4 LOCRC NHW CRC

Figure 1d depicts the RTK-RAS alteration profile in LOCRC NHW colorectal cancer (LOCRC; n= 1,538), integrating mutation class, TMB, and FOLFOX treatment annotation. Overall, 1,108 tumors (72.0%) harbored at least one RTK-RAS pathway alteration, underscoring the pervasive involvement of this signaling axis in LOCRC NHW disease.

Across the cohort, KRAS was the most frequently altered gene (44%), overwhelmingly driven by missense substitutions consistent with activating oncogenic variants. Additional recurrent alterations included BRAF (13%), NF1 (6%), and several receptor tyrosine kinases, most notably ERBB2 (5%), ERBB3 (5%), ERBB4 (5%), ROS1 (5%), and IGF1R (4%). A broad distribution of lower-frequency events (approximately 1-3%) involved NRAS, RET, KIT, FGFR-family members, ALK, MET, ARAF, RAF1, and downstream MAPK regulators, reflecting extensive pathway heterogeneity.

The majority of observed variants were missense (Table S13), with smaller contributions from in-frame insertions/deletions, frameshift events, nonsense mutations, splice-site alterations, and multi-hit configurations, particularly among RTK family members and pathway regulators. TMB values were predominantly low to moderate, with a limited number of tumors showing elevated TMB that did not cluster within specific RTK-RAS genes, suggesting diverse mutational processes rather than a single hypermutated pathway-driven subset.

Assessment of treatment status revealed that FOLFOX-treated and untreated LO NHW tumors were broadly interspersed across genes and mutation classes, with no clear segregation by treatment exposure in the overall RTK-RAS mutational landscape. When considered alongside EOCRC NHW CRC, LOCRC disease exhibited a modestly higher prevalence of pathway alterations and a greater contribution from BRAF and receptor tyrosine kinase alterations, highlighting age-associated shifts within an otherwise conserved RTK-RAS-driven oncogenic framework.

### 3.6 Survival Impact of RTK-RAS Pathway Alterations Across Age, Ancestry, and Treatment Groups

We evaluated the association between RTK-RAS pathway alteration status and overall survival (OS) in EOCRC and LOCRC using Kaplan-Meier analysis, stratifying patients by age at onset, ancestry, and exposure to FOLFOX chemotherapy.

Among EOCRC H/L patients treated with FOLFOX, RTK-RAS pathway alterations were not associated with differences in overall survival (Figure 2a). Survival curves for altered and non-altered tumors were nearly superimposable across the entire follow-up period, with no sustained divergence observed (log-rank p = 0.94). Survival probabilities declined gradually and in parallel for both groups, and the wide, overlapping confidence intervals, particularly at later time points, reflected diminishing numbers at risk rather than a biologically meaningful separation. These findings indicate that RTK-RAS alteration status does not influence OS in FOLFOX-treated early-onset H/L patients.

**Figure 2.**
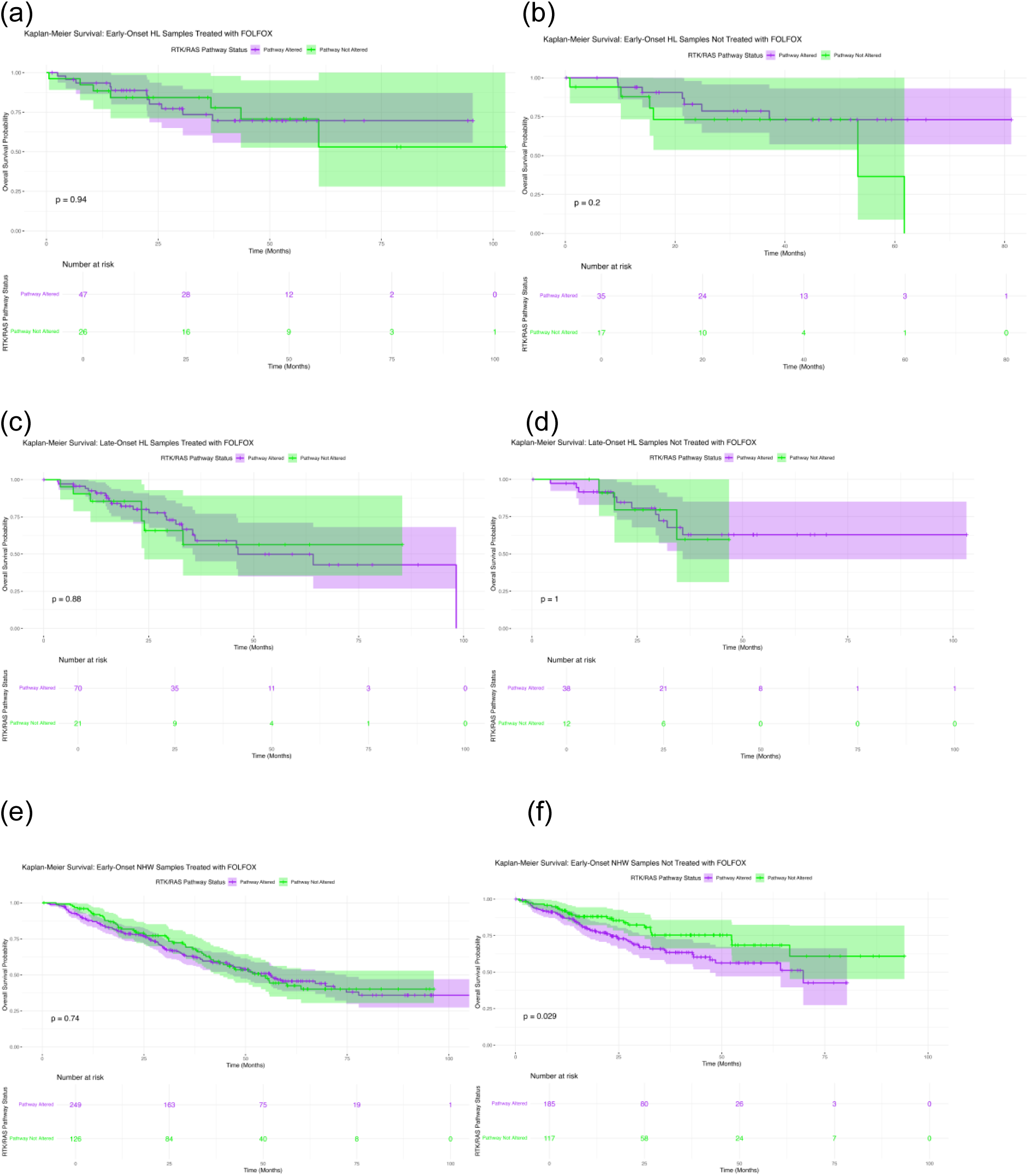
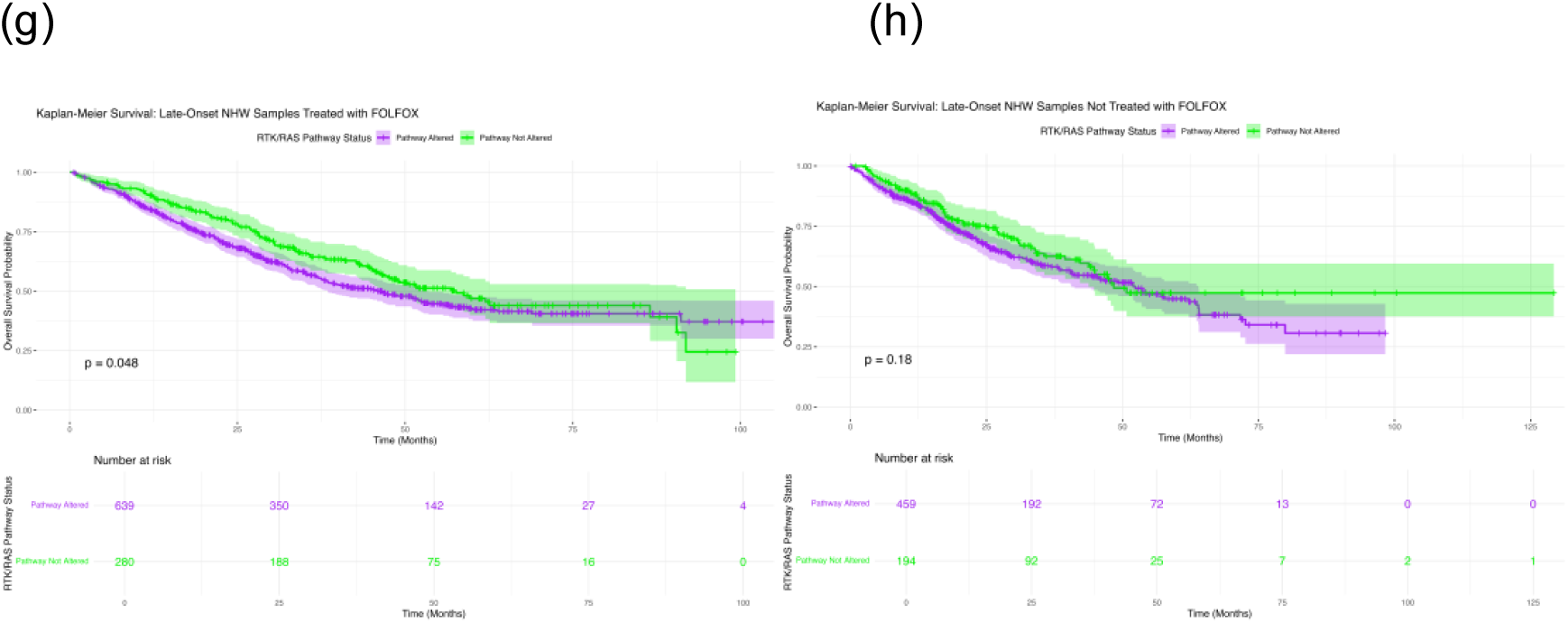
Prognostic significance of RTK-RAS pathway alterations across colorectal cancer subgroups stratified by age at onset, ancestry, and FOLFOX exposure. Kaplan-Meier overall survival curves are presented according to RTK-RAS pathway alteration status (altered vs. non-altered) within clinically and demographically defined CRC subgroups. Panels include: (a-b) early-onset (EOCRC) Hispanic/Latino (H/L) patients treated with or not treated with FOLFOX; (c-d) late-onset (LOCRC) H/L patients treated with or not treated with FOLFOX; (e-f) EOCRC non-Hispanic White (NHW) patients treated with or not treated with FOLFOX; and (g-h) LOCRC NHW patients treated with or not treated with FOLFOX. Shaded regions denote 95% confidence intervals, and numbers at risk are displayed below each plot to facilitate comparison of subgroup-specific survival trajectories associated with RTK-RAS pathway alterations.

Similarly, in EOCRC H/L patients who did not receive FOLFOX, overall survival did not differ by RTK-RAS pathway status (Figure 2b). Kaplan-Meier curves remained largely overlapping throughout follow-up, and no statistically significant association was detected (log-rank p = 0.20). Although a modest separation emerged at later time points, this pattern was driven by a small number of events and was accompanied by broad confidence intervals, limiting interpretability. Collectively, these results suggest that RTK-RAS alterations do not materially affect survival outcomes in early-onset H/L CRC in the absence of oxaliplatin-based chemotherapy.

In LOCRC H/L patients treated with FOLFOX, survival outcomes were likewise independent of RTK-RAS pathway status (Figure 2c). Kaplan-Meier estimates for altered and non-altered tumors closely tracked one another across follow-up, with no significant difference observed (log-rank p = 0.88). Minor early fluctuations between curves were not sustained and were accompanied by overlapping confidence intervals, indicating no consistent prognostic effect.

A comparable pattern was observed in LOCRC H/L patients not treated with FOLFOX (Figure 2d). Survival curves for altered and non-altered tumors were virtually indistinguishable across the full duration of follow-up, and statistical testing confirmed the absence of any survival difference (log-rank p = 1.0). These findings collectively demonstrate that, across all age and treatment strata within the H/L population, RTK-RAS pathway alteration status does not confer prognostic significance for overall survival.

In contrast, ancestry- and treatment-specific effects emerged within the Non-Hispanic White (NHW) cohort. Among early-onset NHW CRC patients treated with FOLFOX, RTK-RAS pathway alterations were not associated with OS differences (Figure 2e). The Kaplan-Meier curves followed closely aligned trajectories, and formal comparison showed no significant association (log-rank p = 0.74). Minor divergence at intermediate time points was accompanied by extensive overlap of confidence intervals, suggesting that these fluctuations do not reflect a robust biological effect.

However, among EOCRC NHW patients who did not receive FOLFOX, RTK-RAS pathway alteration status was significantly associated with inferior overall survival (Figure 2f). Patients with RTK-RAS-altered tumors exhibited a clear and persistent survival disadvantage compared with non-altered patients, as evidenced by early separation of Kaplan-Meier curves that widened over time (log-rank p = 0.029). Although confidence intervals broadened at later follow-up due to decreasing numbers at risk, the sustained divergence supports a negative prognostic impact of RTK-RAS alterations in untreated early-onset NHW CRC.

Conversely, in LOCRC NHW patients treated with FOLFOX, RTK-RAS pathway alterations were associated with improved overall survival (Figure 2g). Altered tumors demonstrated consistently higher survival probabilities compared with non-altered tumors, with progressive separation of Kaplan-Meier curves over follow-up (log-rank p = 0.048). While confidence intervals widened toward the tail of follow-up, the sustained and directional curve separation suggests a favorable prognostic association of RTK-RAS alterations in this treatment-defined subgroup, contrasting sharply with the adverse effect observed in untreated early-onset NHW disease.

Finally, among LOCRC NHW patients who did not receive FOLFOX, RTK-RAS pathway status was not significantly associated with OS (Figure 2h). Survival curves for altered and non-altered tumors followed largely parallel trajectories, with only modest late separation and substantial overlap of confidence intervals (log-rank p = 0.18). Although non-altered patients appeared to maintain slightly higher long-term survival, this trend did not reach statistical significance, indicating no clear prognostic effect in this subgroup.

These analyses reveal a striking age-, ancestry-, and treatment-dependent relationship between RTK-RAS pathway alterations and survival. RTK-RAS alterations conferred adverse prognosis in untreated early-onset NHW CRC, were associated with improved survival in FOLFOX-treated late-onset NHW CRC, and showed no measurable survival impact across all Hispanic/Latino subgroups, regardless of age or treatment exposure. These findings highlight the context-dependent nature of RTK-RAS pathway biology and underscore the importance of integrating ancestry, age at onset, and chemotherapy exposure when interpreting genomic prognostic biomarkers in CRC.

### 3.7 AI-driven exploratory interrogation and hypothesis refinement

To enable efficient hypothesis generation prior to formal statistical testing, the AI-HOPE (38) and AI-HOPE-RTK-RAS (39) platforms were applied to conduct an automated, post hoc exploration of the integrated colorectal cancer (CRC) dataset. Through natural language-based querying of harmonized clinical, molecular, and treatment-related variables, the AI framework rapidly surfaced candidate associations with potential biological and clinical relevance, which were subsequently evaluated using confirmatory statistical methods.

As an initial finding, AI-guided interrogation identified a possible survival disadvantage associated with RTK-RAS pathway alterations in EOCRC NHW CRC patients who had not received FOLFOX chemotherapy. To formally assess this observation, a targeted case-control analysis was performed, defining cases as EOCRC NHW patients harboring at least one RTK-RAS pathway alteration (n = 185) and controls as EOCRC NHW patients without such alterations (n = 117), all without FOLFOX exposure. Kaplan-Meier survival analysis demonstrated significantly inferior overall survival among patients with RTK-RAS-altered tumors compared with their non-altered counterparts (log-rank p = 0.0288) (Figure S1). Notably, survival curves diverged early in follow-up, with altered cases experiencing a more rapid decline in survival probability, supporting a potential prognostic role for RTK-RAS pathway dysregulation independent of oxaliplatin-based treatment.

Next, the AI platform executed a structured evaluation of cohort composition under tightly defined clinical and genomic constraints to determine whether preliminary pathway enrichment signals persisted across EOCRC CRC subgroups. This analysis revealed that, for both case and control cohorts, samples satisfying the analytic context constituted a small and highly specific subset of the overall dataset, underscoring the precision of AI-guided cohort selection and minimal inclusion of out-of-context samples. Subsequent Fisher’s exact testing did not demonstrate significant enrichment of the queried genomic feature between the compared groups (Figure S2). These findings illustrate how AI-facilitated exploratory signals can be rigorously examined through transparent cohort construction and pre-statistical validation, enabling discrimination between true biological patterns and chance findings.

In parallel, AI-enabled global scanning identified a broad set of clinical and molecular features that differed significantly between EOCRC H/L and EOCRC NHW CRC patients, regardless of treatment status (p < 0.05). The identified variables spanned demographic characteristics (ethnicity and race), diagnostic and tumor features (cancer type, diagnosis description, sample type, and stage at diagnosis), as well as clinical outcomes (overall survival status and event occurrence). Additionally, several key driver genes, including SMAD4, APC, TP53, KRAS, and BRAF, exhibited differential mutation prevalence across ancestries (Figure S3). Tumor anatomical site and stage at diagnosis emerged as particularly prominent distinguishing factors, aligning with prior reports of ancestry-associated variation in EOCRC CRC presentation. Together, these results highlight the capacity of AI-guided interrogation to uncover multidimensional heterogeneity across populations and to systematically prioritize variables for downstream analyses.

Finally, AI-driven screening highlighted a potential ancestry-related difference in BRAF mutation frequency among EOCRC CRC patients treated with FOLFOX. This observation prompted a focused case-control comparison between EOCRC H/L patients (n = 73) and EOCRC NHW patients (n = 375) who received FOLFOX chemotherapy. Statistical evaluation using Fisher’s exact test revealed a differential distribution of BRAF mutations between the two ancestry groups (Figure S4). Although BRAF alterations were infrequent overall, the observed imbalance suggests possible ancestry-associated differences in RTK-RAS pathway architecture within this clinically relevant treatment subgroup, meriting further biological and therapeutic investigation.

In summary, these AI-derived exploratory analyses informed the selection of clinically meaningful subgroup comparisons and guided the prioritization of variables for formal statistical testing. By automating cohort definition, filtering, and visualization across complex clinical and genomic dimensions, the AI-HOPE (38) framework reduced manual analytic burden, improved reproducibility, and streamlined the progression from hypothesis generation to confirmatory precision oncology analysis.

## 4. Discussion

In this study, we applied an AI-enabled precision oncology workflow, AI-HOPE(38) and the pathway-specialized AI-HOPE-RTK-RAS agent (39), to interrogate RTK-RAS alterations in colorectal cancer (CRC) at the intersection of age at onset, ancestry, and FOLFOX exposure. By integrating harmonized clinical annotations with pathway-level somatic mutation calls across 2,515 cases, we identified context-dependent genomic patterns and divergent survival associations that would be difficult to resolve without high-dimensional subgrouping. Collectively, our results support a model in which RTK-RAS dysregulation is common across CRC, yet its clinical meaning varies substantially depending on treatment exposure and patient subgroup.

### RTK-RAS alterations show treatment- and age-dependent prognostic polarity in NHW CRC

A central observation was that RTK-RAS pathway status was not uniformly prognostic across the cohort. Instead, survival associations emerged only in specific strata. In EOCRC NHW patients not treated with FOLFOX, RTK-RAS alterations were associated with inferior overall survival (log-rank p ≈ 0.029). This suggests that, in the absence of oxaliplatin-based therapy, RTK-RAS pathway disruption may act as a surrogate for more aggressive biology, potentially reflecting higher baseline signaling flux through RAS/RAF/MAPK, greater metastatic competence, or enrichment for adverse molecular co-alteration profiles that were not explicitly modeled here.

In contrast, in LOCRC NHW patients treated with FOLFOX, RTK-RAS alterations were associated with improved overall survival (p ≈ 0.048), indicating an opposite directional effect. One biologically plausible explanation is that, in LO disease, certain RTK-RAS-altered tumors may remain more chemosensitive or may represent a subgroup with distinct clinical trajectories (e.g., differential metastatic patterns, treatment sequencing, or tumor burden at therapy initiation). Alternatively, RTK-RAS alteration status may track with a broader molecular state that interacts favorably with fluoropyrimidine/oxaliplatin exposure in older NHW patients. Regardless of mechanism, this polarity highlights an important principle: pathway alteration status can be prognostic in one context and neutral, or even favorable, in another, reinforcing the need for subgroup-aware biomarker interpretation.

### Gene-level differences suggest selection pressures linked to FOLFOX exposure

Although overall RTK-RAS pathway alteration prevalence was high across strata, gene-level analyses revealed selective patterns consistent with treatment-associated pressures or confounding by treatment selection. In EO H/L, ERBB2 and NF1 alterations were less frequent among FOLFOX-treated versus untreated patients, and in LO H/L, NTRK2 alterations were depleted in FOLFOX-exposed cases. In NHW, the most prominent treatment-associated depletion occurred in LO NHW, where multiple RTK-RAS genes (e.g., ERBB3, KIT, IGF1R, RET, ALK, FLT3, ERRFI1, ARAF, RAF1) were less enriched among FOLFOX-treated patients. These consistent reductions across multiple loci suggest that FOLFOX-exposed subgroups may differ systematically from unexposed groups, either because treatment selection correlates with clinical variables (stage, resectability, comorbidities, metastatic site), or because particular molecular contexts are underrepresented among FOLFOX-treated patients due to prior therapy, lineage effects, or survivorship bias. Future analyses incorporating stage-stratified models, regimen timing, and metastatic status will help disentangle true gene-treatment interactions from treatment selection effects.

### Ancestry-associated patterns are most evident in untreated EOCRC disease

Between-ancestry comparisons indicated that ancestry-linked differences were not uniform across therapy strata. Notably, among EO patients not treated with FOLFOX, H/L patients showed enrichment of select RTK-RAS genes (including ERBB2, MAPK3, CBL, and NF1) relative to NHW. This pattern was less apparent in FOLFOX-treated EO patients, suggesting that chemotherapy exposure (or the clinical scenarios that lead to exposure) may partially obscure or reshape detectable ancestry-linked differences. These observations align with the broader view that ancestry-associated disparities in CRC are likely multi-factorial, reflecting tumor biology, host factors, environmental exposures, and care pathways, such that ancestry signals may be most visible in strata with fewer treatment-related filters.

### Implications for precision oncology: RTK-RAS status may inform subgroup-specific risk and therapy interpretation

From a clinical standpoint, our findings argue against using a single “RTK-RAS altered” label as a universal prognostic biomarker across CRC. Instead, RTK-RAS pathway alterations may serve as subgroup-specific markers that modify risk in a way that depends on age and treatment context. The adverse survival association in untreated EO NHW disease suggests a potential use case for RTK-RAS status in identifying higher-risk patients who might benefit from intensified surveillance or alternative systemic strategies, while the favorable association in FOLFOX-treated LO NHW disease raises the possibility that, in some older patients, RTK-RAS alterations track with tumors that respond better to standard chemotherapy. These hypotheses require validation in independent cohorts and, ideally, in analyses that incorporate additional clinical covariates, lines of therapy, and metastatic setting.

### AI-HOPE as an enabling framework for subgroup-aware biomarker discovery

A key contribution of this work is methodological: the AI-HOPE (38) / AI-HOPE-RTK-RAS (39) agents accelerated cohort construction, stratified querying, and rapid testing of clinically meaningful hypotheses across numerous intersections of ancestry, age, and treatment. Importantly, the AI-driven workflow did not replace statistical inference; rather, it functioned as a scalable discovery layer that identifies candidate signals, constructs transparent case-control subsets, and prioritizes comparisons for confirmatory testing. This approach is particularly relevant for EOCRC and health-equity research, where meaningful subgrouping can quickly lead to sparse strata and complex multi-parameter definitions that are cumbersome to operationalize manually.

### Limitations and future directions

Several limitations should be considered. First, although the overall cohort was large, certain subgroup analyses, especially when stratifying by ancestry, age, and treatment, reduced effective sample sizes, which can widen confidence intervals and increase susceptibility to instability. Second, FOLFOX exposure was treated as a binary annotation without granular details (line of therapy, dosing intensity, duration, adjuvant vs metastatic use, and sequencing with targeted agents), limiting causal interpretation of treatment interactions. Third, our RTK-RAS definition relied on protein-altering mutations and did not incorporate other alteration classes (copy-number changes, fusions, or expression-based activation), which may be important for RTK biology. Finally, because these were retrospective, multi-cohort public datasets, residual confounding by clinical covariates and ascertainment differences is unavoidable.

Future work should (i) validate these subgroup-specific survival associations in independent ancestrally diverse cohorts; (ii) incorporate richer treatment timelines and metastatic context; (iii) extend RTK-RAS alteration calls to include CNAs/fusions where available; and (iv) integrate microenvironmental and functional context using emerging modalities such as spatial profiling and single-cell analyses to clarify mechanisms linking RTK-RAS dysregulation to chemotherapy response and survival.

Overall, this AI-enabled analysis demonstrates that RTK-RAS pathway alterations carry strong context dependency, with survival effects that differ by age at onset, ancestry, and chemotherapy exposure. These findings support a precision oncology approach in which pathway biomarkers are interpreted within explicit clinical strata, and they highlight the value of AI-assisted analytic frameworks for scalable, subgroup-aware biomarker discovery in populations affected by EOCRC.

## Data Availability

All data used in the present study is publicly available at https://www.cbioportal.org/ and https://genie.cbioportal.org. The datasets used in our study were aggregated/summary data, and no individual-level data were used. Additional data can be provided upon reasonable request to the authors.

## Supplementary Materials

**Table S1.**
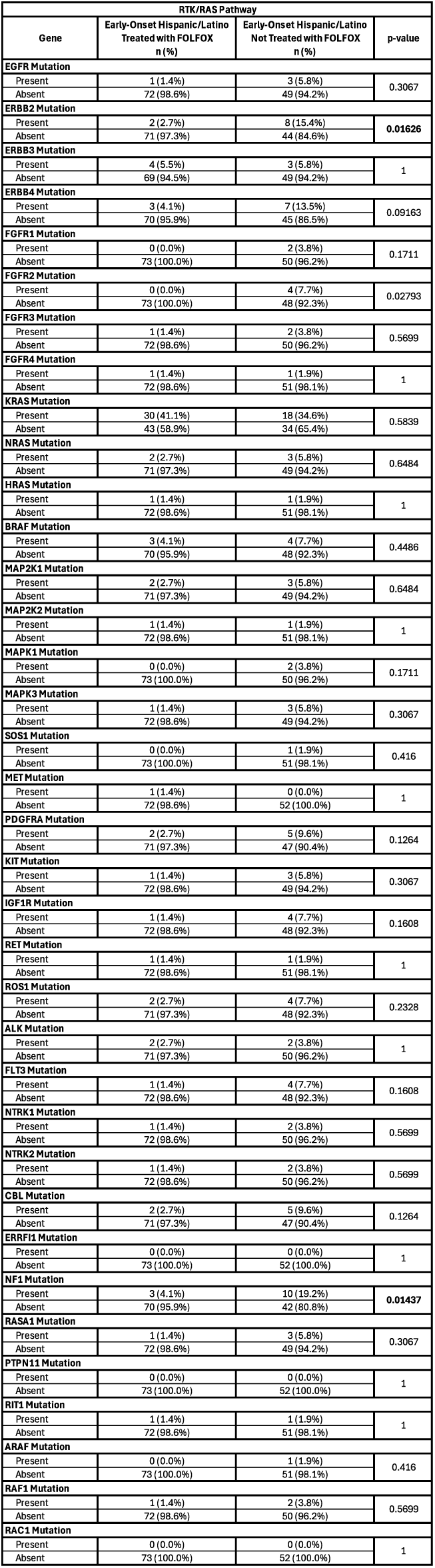
Comparison of RTK-RAS pathway alteration frequencies in early-onset Hispanic/Latino (H/L) colorectal cancer patients treated with FOLFOX versus not treated with FOLFOX.

**Table S2.**
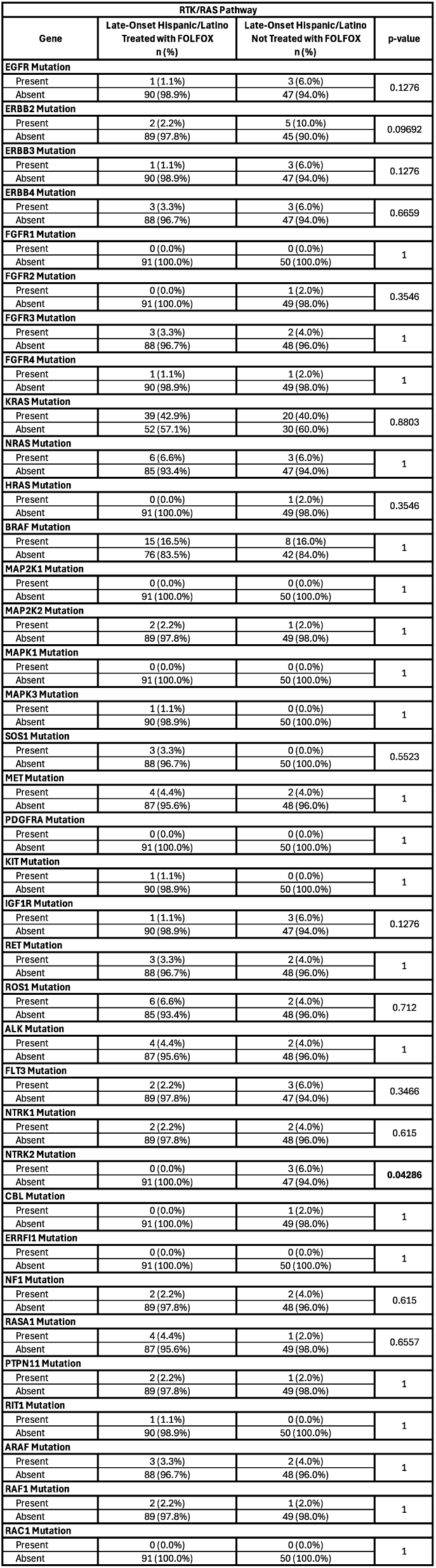
Comparison of RTK-RAS pathway alteration frequencies in late-onset Hispanic/Latino (H/L) colorectal cancer patients treated with FOLFOX versus not treated with FOLFOX.

**Table S3.**
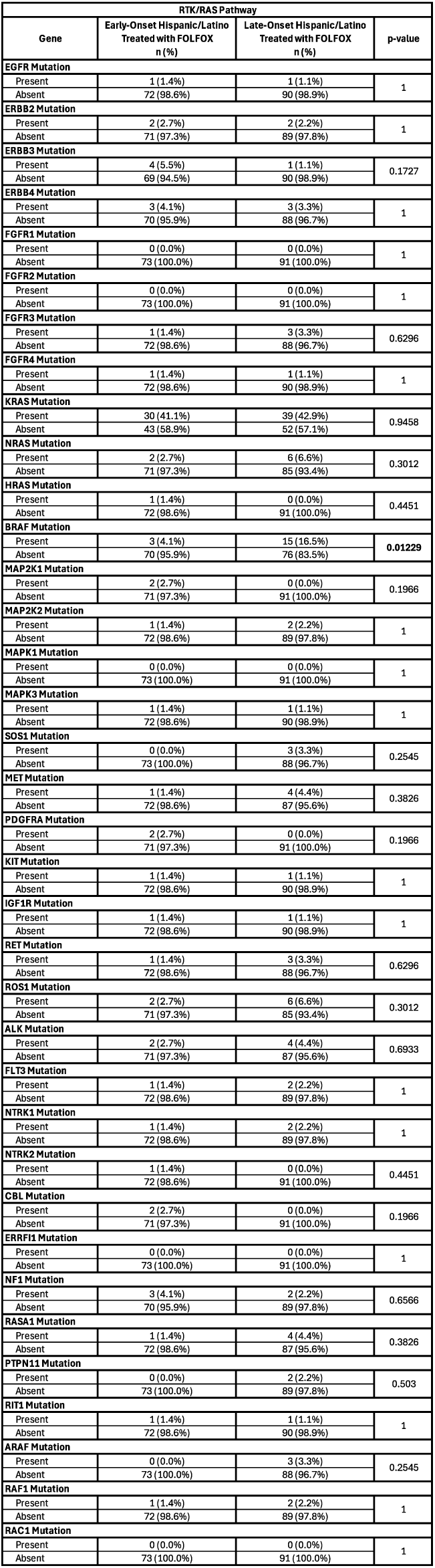
Comparison of RTK-RAS pathway alteration frequencies between early-onset and late-onset Hispanic/Latino (H/L) colorectal cancer patients treated with FOLFOX.

**Table S4.**
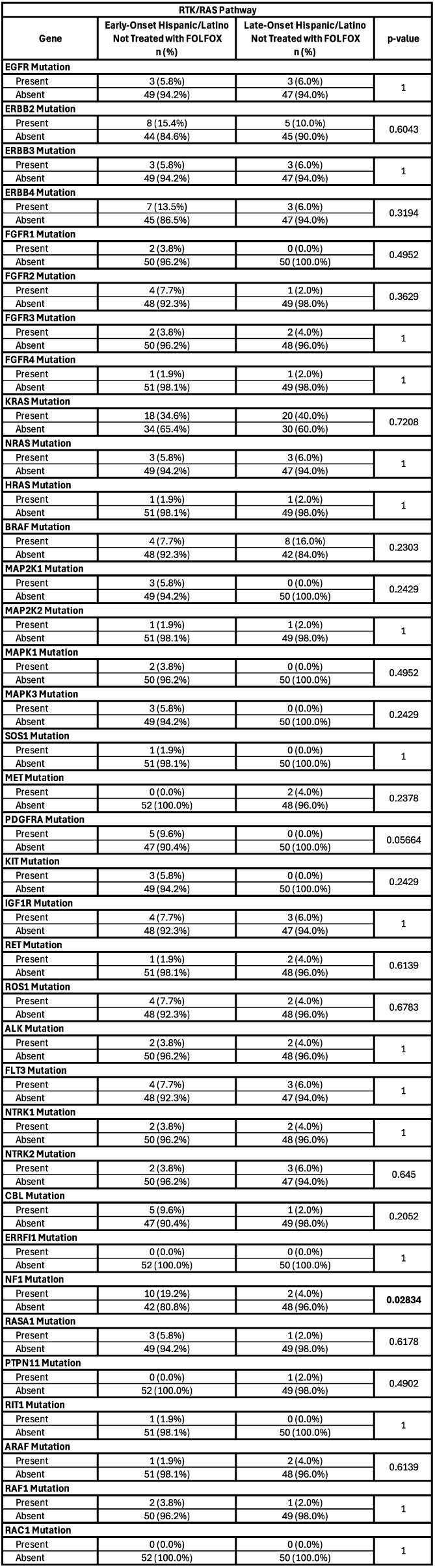
Comparison of RTK-RAS pathway alteration frequencies between early-onset and late-onset Hispanic/Latino (H/L) colorectal cancer patients not treated with FOLFOX.

**Table S5.**
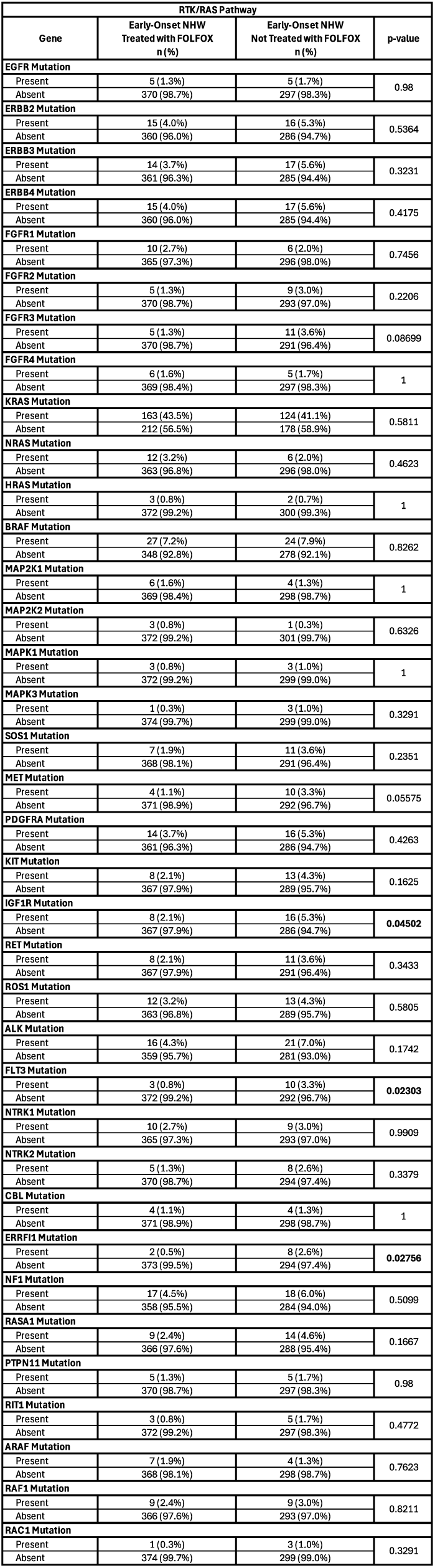
Comparison of RTK-RAS pathway alteration frequencies in early-onset Non-Hispanic White (NHW) colorectal cancer patients treated with FOLFOX versus not treated with FOLFOX.

**Table S6.**
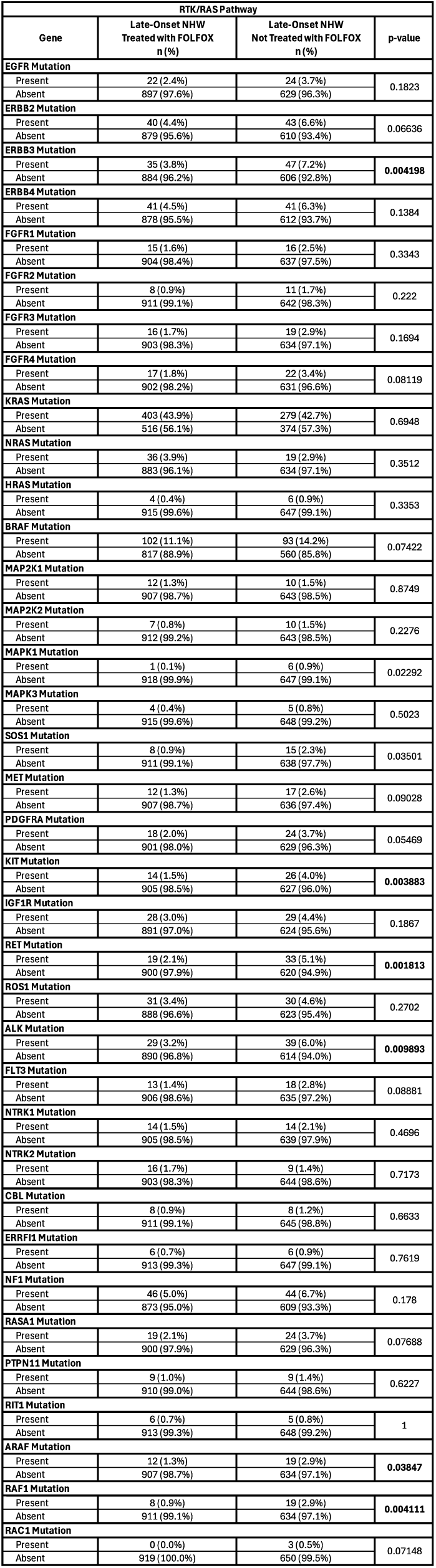
Comparison of RTK-RAS pathway alteration frequencies in late-onset Non-Hispanic White (NHW) colorectal cancer patients treated with FOLFOX versus not treated with FOLFOX.

**Table S7.**
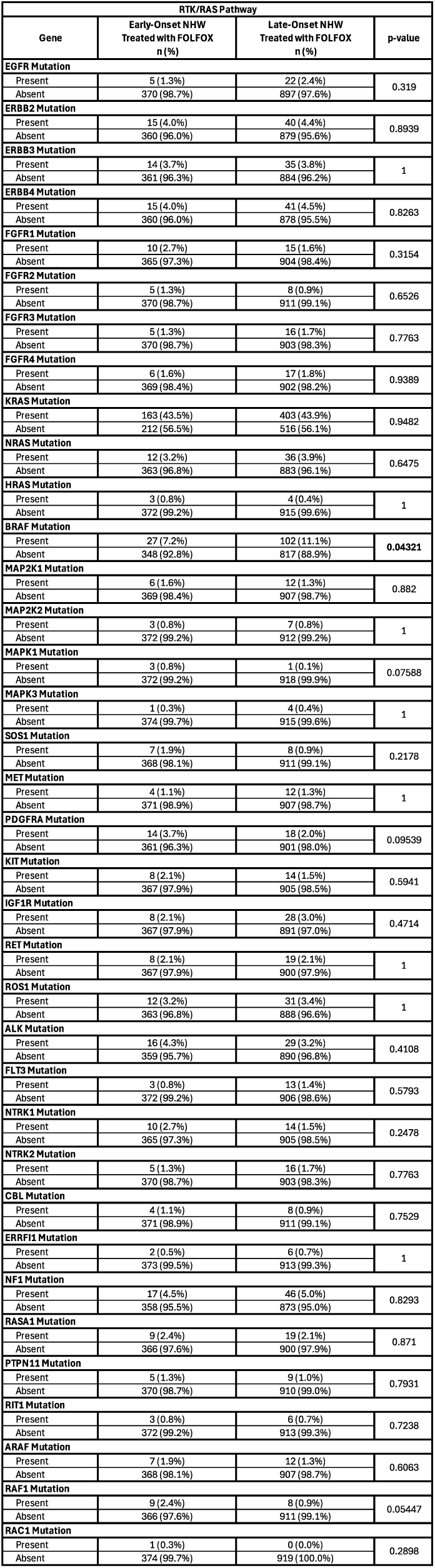
Comparison of RTK-RAS pathway alteration frequencies between early-onset and late-onset Non-Hispanic White (NHW) colorectal cancer patients treated with FOLFOX.

**Table S8.**
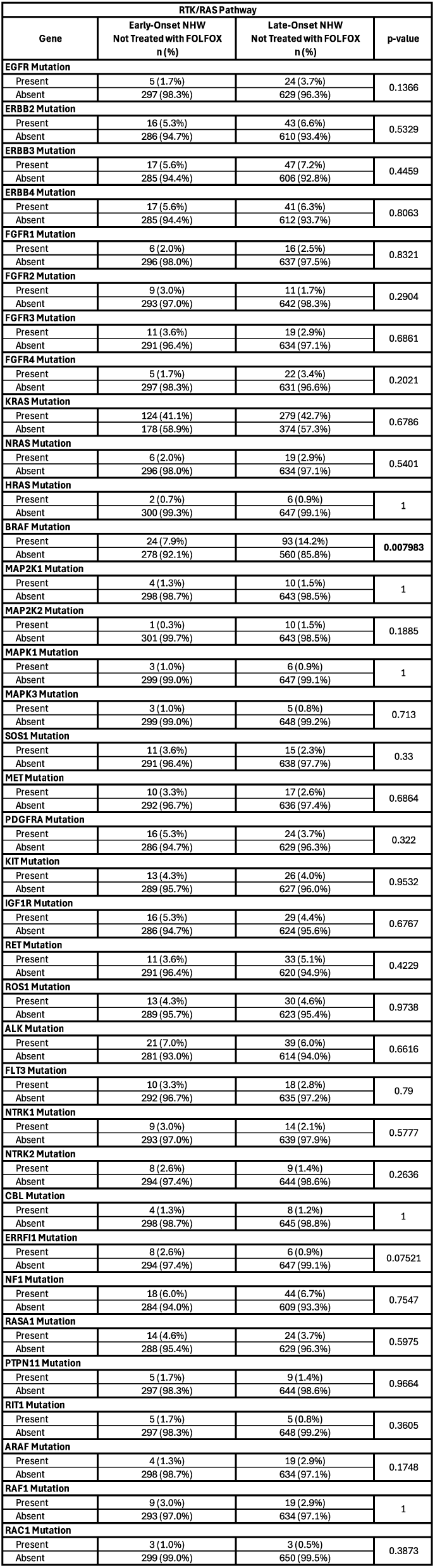
Comparison of RTK-RAS pathway alteration frequencies between early-onset and late-onset Non-Hispanic White (NHW) colorectal cancer patients not treated with FOLFOX.

**Table S9.**
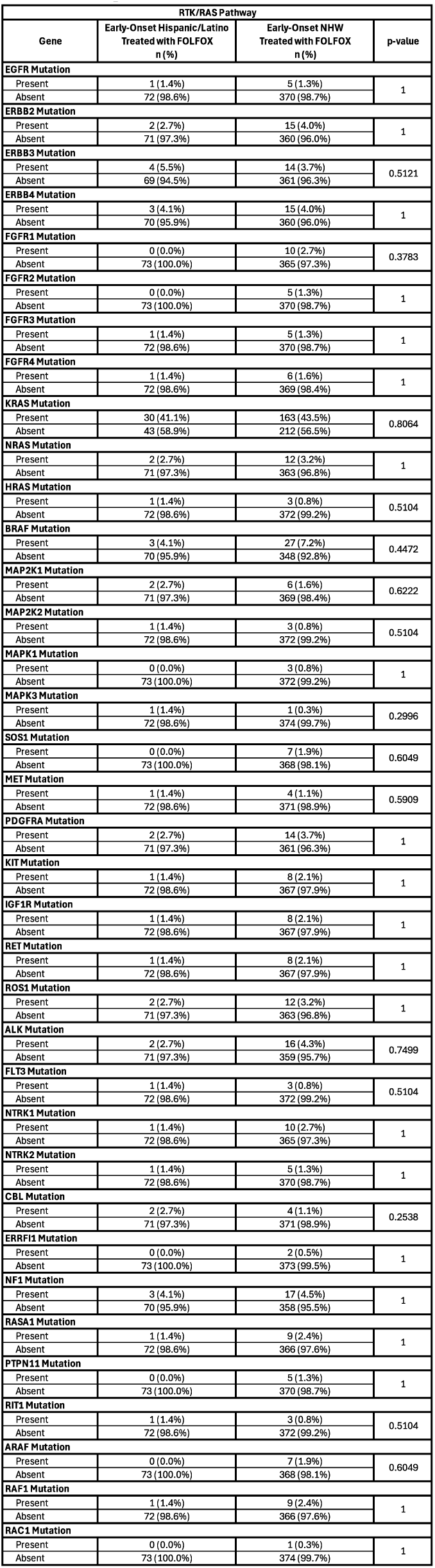
Comparison of RTK-RAS pathway alteration frequencies between early-onset Hispanic/Latino (H/L) and early-onset Non-Hispanic White (NHW) colorectal cancer patients treated with FOLFOX.

**Table S10.**
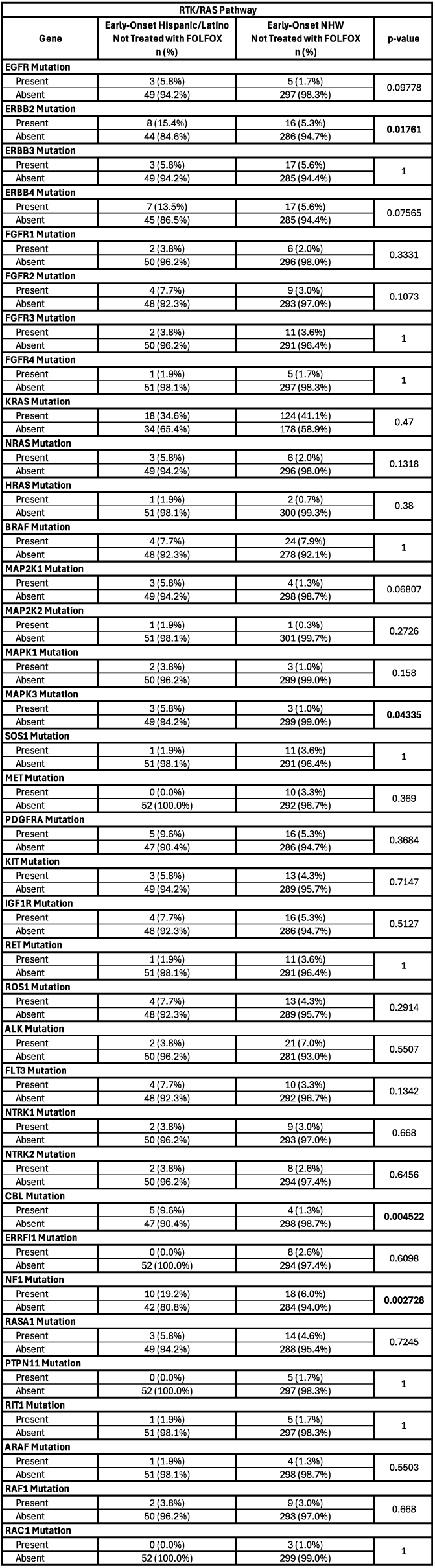
Comparison of RTK-RAS pathway alteration frequencies between early-onset Hispanic/Latino (H/L) and early-onset Non-Hispanic White (NHW) colorectal cancer patients not treated with FOLFOX.

**Table S11.**
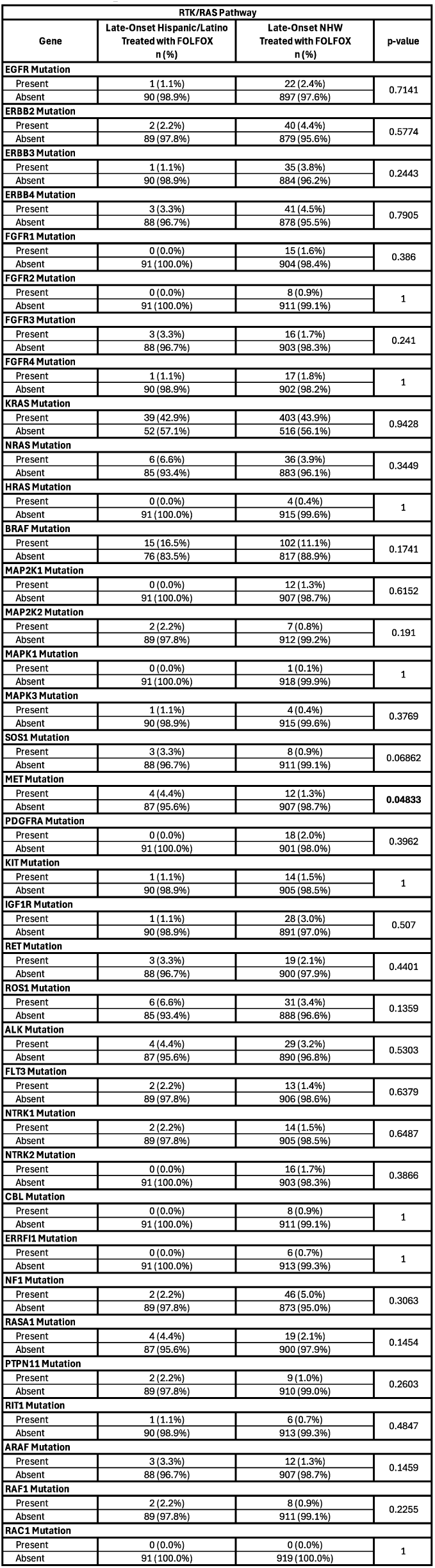
Comparison of RTK-RAS pathway alteration frequencies between late-onset Hispanic/Latino (H/L) and late-onset Non-Hispanic White (NHW) colorectal cancer patients treated with FOLFOX.

**Table S12.**
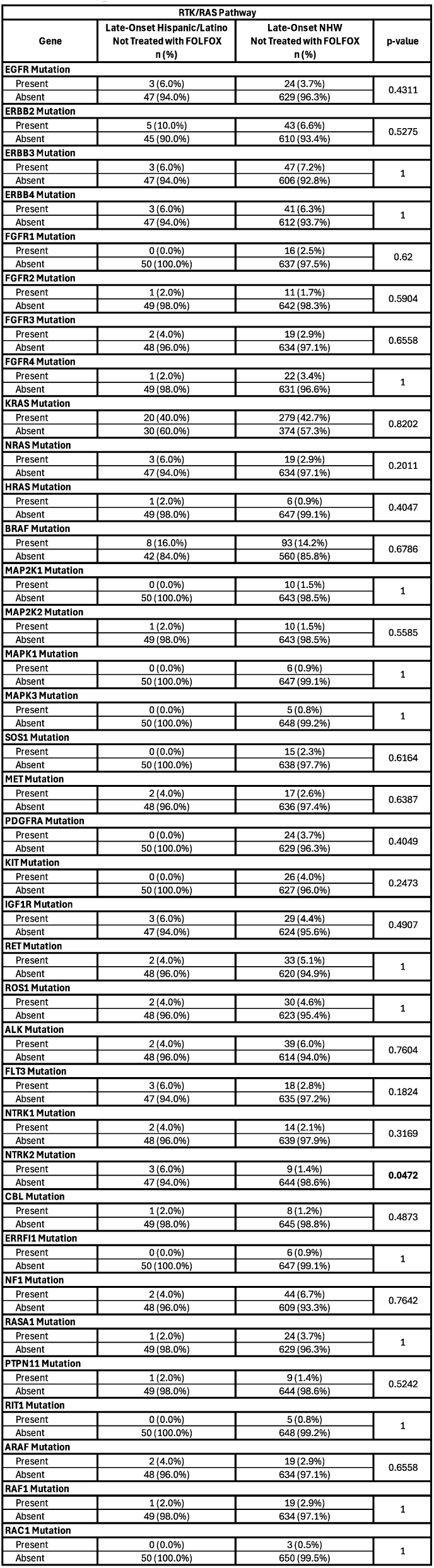
Comparison of RTK-RAS pathway alteration frequencies between late-onset Hispanic/Latino (H/L) and late-onset Non-Hispanic White (NHW) colorectal cancer patients not treated with FOLFOX.

**Table S13.**
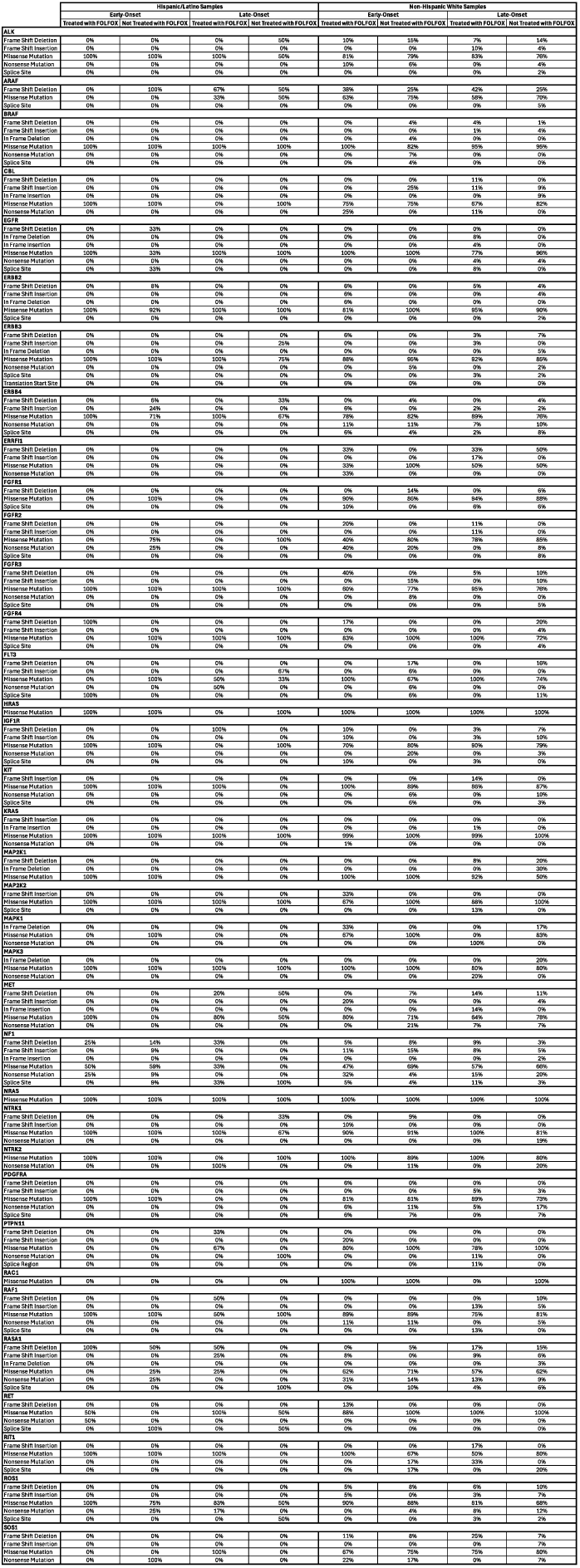
Spectrum of RTK-RAS pathway variant classes across colorectal cancer subgroups defined by ancestry, age at diagnosis, and FOLFOX exposure. This table summarizes the relative contribution of distinct somatic variant classes observed in selected RTK-RAS pathway genes, stratified by Hispanic/Latino (H/L) versus non-Hispanic White (NHW) ancestry, early-onset (EOCRC) versus late-onset (LOCRC) disease, and receipt of FOLFOX chemotherapy. Variant categories include truncating, non-truncating, splice-associated, and translation-initiation-related alterations, encompassing both insertion/deletion and single-nucleotide change events. Values are expressed as percentages representing the distribution of each variant class among all detected alterations for a given gene within each subgroup. This stratified presentation enables comparison of mutation pattern heterogeneity across demographic and treatment-defined CRC populations and provides insight into potential differences in underlying mutational processes.

**Figure S1.**
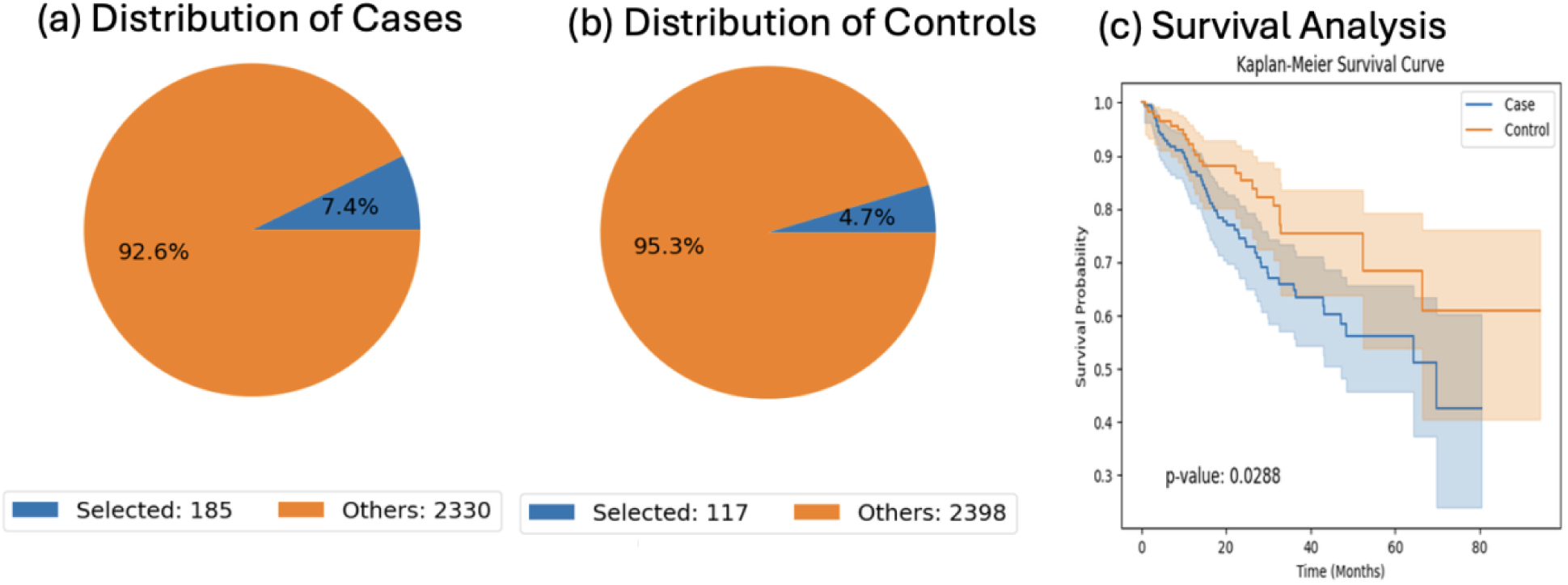
AI-enabled cohort definition and survival stratification of early-onset colorectal cancer (EOCRC) non-Hispanic White (NHW) patients not treated with FOLFOX based on RTK-RAS pathway status. AI-HOPE was used to programmatically define case and control cohorts using integrated clinical variables (ancestry, age at onset, treatment exposure) and pathway-level genomic annotations. The case cohort consisted of EOCRC NHW patients who did not receive FOLFOX and harbored at least one RTK-RAS pathway alteration (n = 185), while the control cohort included EOCRC NHW patients without RTK-RAS alterations and no FOLFOX exposure (n = 117). Pie charts illustrate the proportion of selected samples relative to the full dataset for each cohort. Kaplan-Meier analysis demonstrates a significant difference in overall survival between altered and non-altered groups, with RTK-RAS-altered tumors exhibiting inferior survival outcomes (log-rank p = 0.0288). Shaded regions represent 95% confidence intervals, highlighting increased early divergence of survival probabilities between groups.

**Figure S2.**
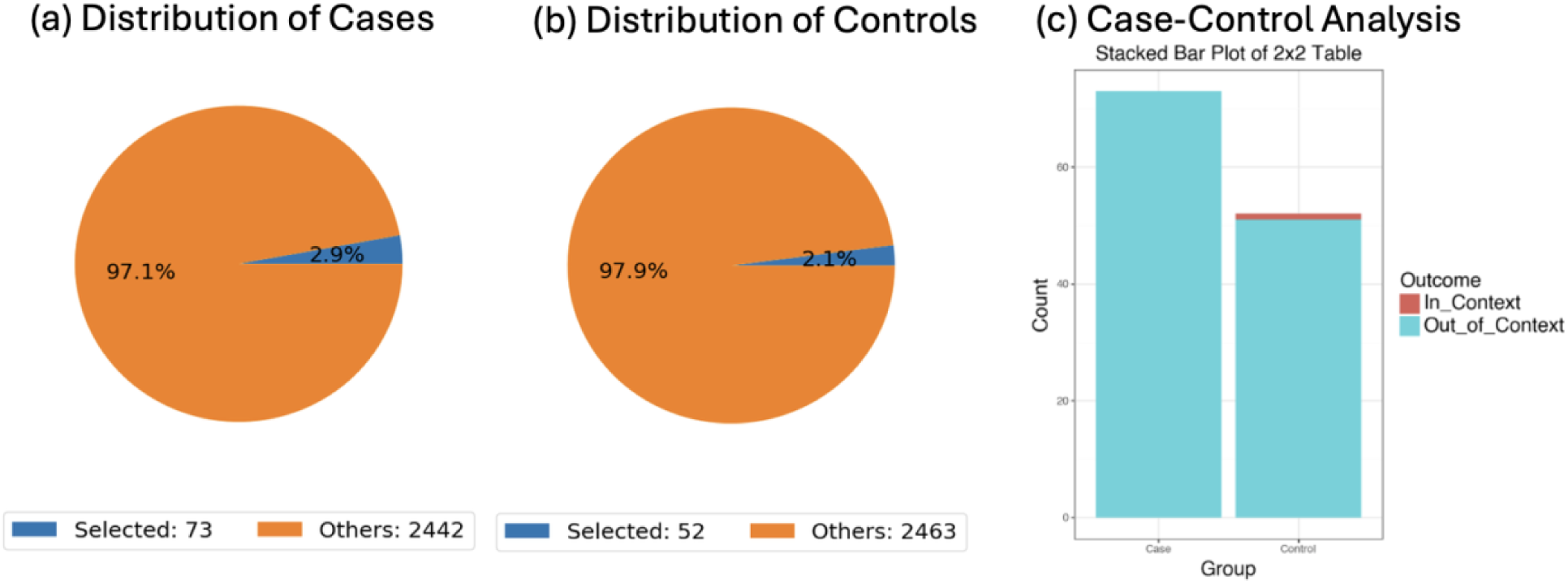
AI-driven case-control composition and comparative analysis of pathway-defined cohorts in early-onset colorectal cancer (EOCRC). This figure summarizes the AI-HOPE-guided construction and evaluation of case and control cohorts derived from EOCRC patients under predefined clinical and genomic constraints. Panel (a) illustrates the proportion of samples meeting the case criteria (selected/in-context) relative to the full dataset, while panel (b) shows the corresponding distribution for the control cohort. In both groups, selected samples represent a small, highly specific subset of the overall population, reflecting stringent cohort definition. Panel (c) presents a stacked bar visualization of the resulting 2×2 case-control table, comparing in-context versus out-of-context samples across groups. Statistical assessment was performed using Fisher’s exact testing to evaluate differences between cohorts, demonstrating no significant enrichment of the queried genomic feature between cases and controls. Together, these plots highlight the transparency of AI-assisted cohort selection and provide a quantitative framework for downstream comparative analyses.

**Figure S3.**
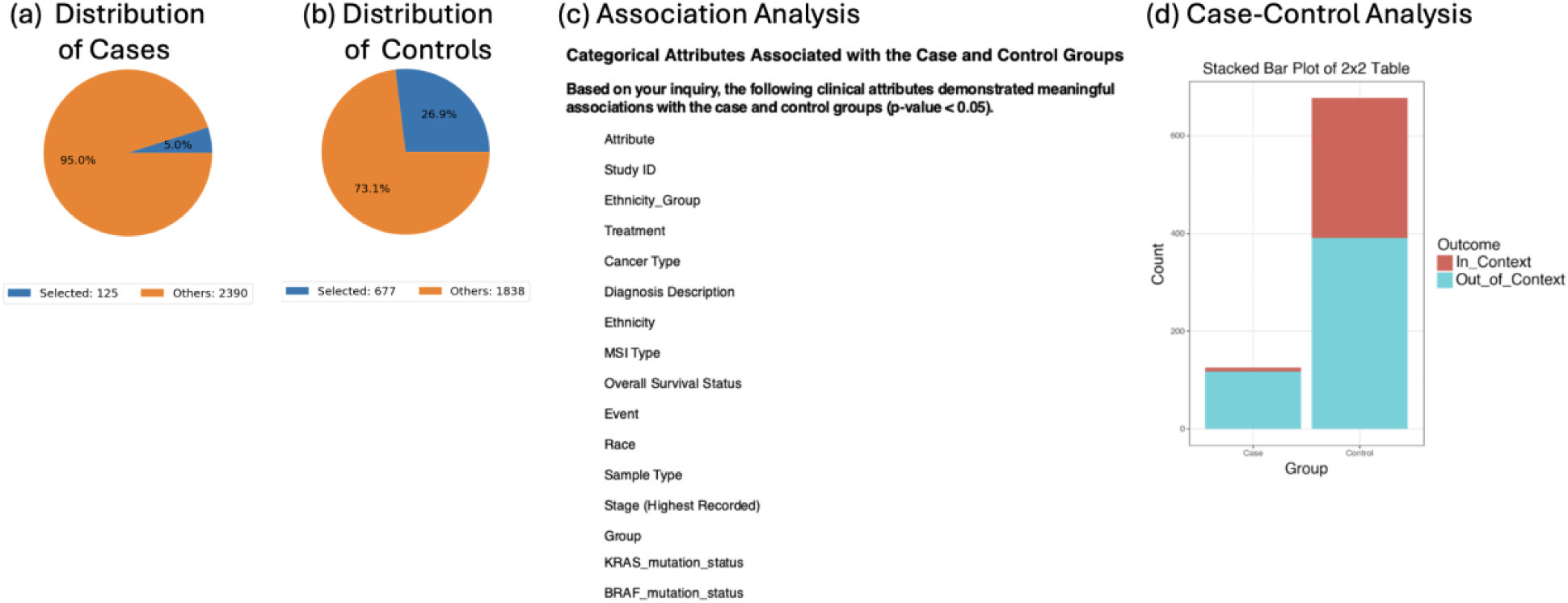
Systematic AI-assisted comparison of clinical and genomic features between early-onset colorectal cancer (EOCRC) Hispanic/Latino (H/L) and Non-Hispanic White (NHW) cohorts. This figure summarizes the output of an AI-HOPE-driven exploratory analysis designed to identify clinical and molecular attributes that distinguish EOCRC cases by ancestry. a) The case cohort comprised EOCRC H/L patients (n = 125), b) while the control cohort included EOCRC NHW patients (n = 677), both defined using uniform age-of-onset criteria. Pie charts depict the relative proportions of samples meeting the user-defined analytic context versus the remaining dataset for each cohort, illustrating the differential enrichment of in-context samples across ancestries. c) The accompanying table enumerates categorical variables demonstrating statistically significant associations between case and control groups (p < 0.05), spanning demographic descriptors (ethnicity and race), diagnostic and tumor characteristics (cancer type, diagnosis description, sample type, and stage at diagnosis), clinical outcomes (overall survival status and event), and recurrent oncogenic alterations, including SMAD4, APC, TP53, and KRAS mutation status. The stacked bar plot provides a visual summary of the 2×2 comparison underlying the odds-ratio framework used to assess group-level enrichment. These results underscore ancestry-associated heterogeneity across multiple clinical and molecular dimensions in EOCRC

**Figure S4.**
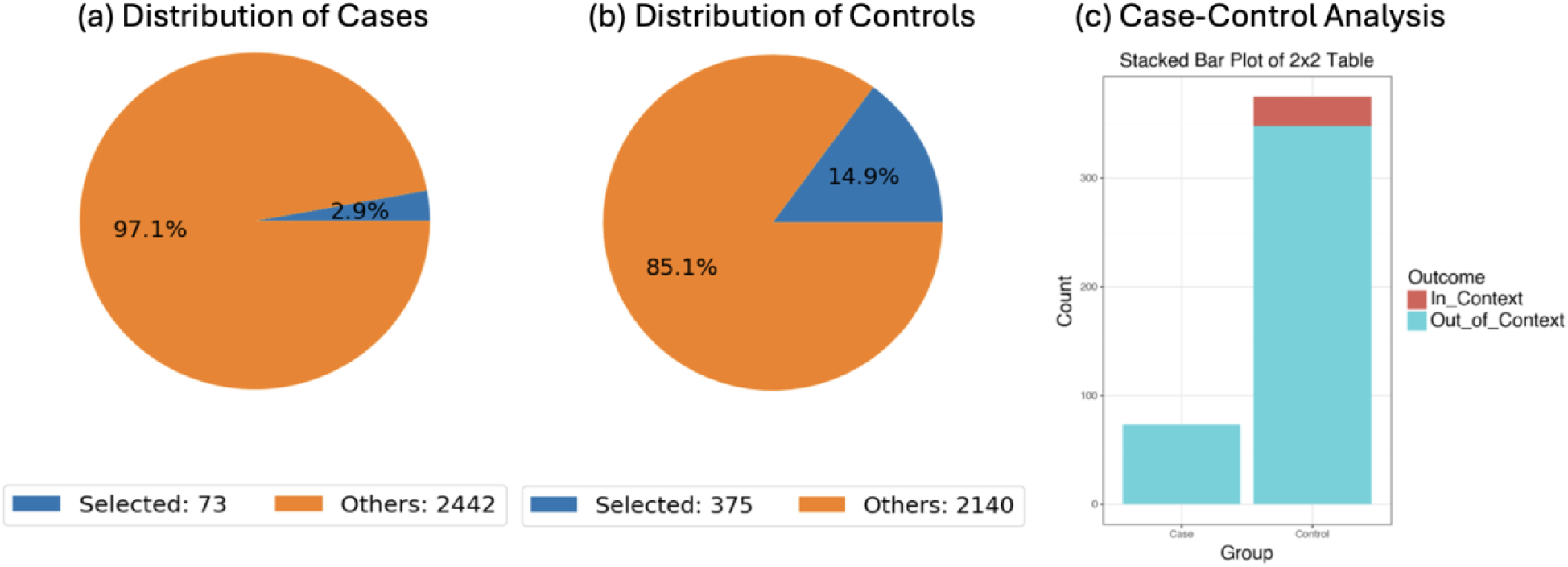
AI-assisted case-control evaluation of BRAF mutation prevalence in FOLFOX-treated EOCRC by ancestry. This figure illustrates an AI-HOPE-guided comparison of BRAF mutation status between EOCRC Hispanic/Latino (H/L) and Non-Hispanic White (NHW) colorectal cancer (CRC) patients who received FOLFOX chemotherapy. The case cohort comprises EOCRC H/L patients (n = 73), while the control cohort includes EOCRC NHW patients (n = 375). (a) and (b) Pie charts depict the proportion of in-context (BRAF-mutant) and out-of-context samples within the case and control cohorts, respectively, highlighting differences in BRAF mutation representation across ancestries. (c) A stacked bar plot summarizes the corresponding 2×2 contingency table used for odds-ratio testing, contrasting BRAF-mutant and wild-type samples between groups. Statistical assessment was performed using Fisher’s exact test, with results indicating a differential distribution of BRAF mutations between EOCRC H/L and NHW patients treated with FOLFOX. Together, these analyses support ancestry-associated variation in RTK-RAS pathway alterations within this clinically relevant treatment subgroup.

